# Molecular Diagnosis as a Probability of Necessity

**DOI:** 10.64898/2026.09.09.26362174

**Authors:** JW Belmont, CJ Williams, CA Shaw

## Abstract

Genetic diagnosis requires attributing a patient’s disease to variant(s). We formalize this attribution through the Probability of Necessity (PN), a counterfactual estimand modeling the probability that disease would not have occurred absent a germline variant given that both are observed, and we use PN to examine the logic and limitations of genetic diagnosis. PN is a property of a variant in a population and disease context, not of a variant alone. PN dissociates from penetrance: identical penetrance yields different PN depending on baseline disease risk and competing causes. Case selection biases PN estimation, favoring population-scale cohorts for estimation over ascertained case series. PN falls as the prevalence of competing causes rises, and among carriers it can fall with increasing polygenic liability even as penetrance rises. We adopt the mediated probability of causation as the Probability of Mediated Necessity (PMN), which updates PN using a biomarker proxying the variant’s operative pathway. Using UK Biobank exome data, we estimate PN and PMN for rare *LDLR* variants in ischemic heart disease (IHD, ICD-10 I25), with hyperlipidemia (E78) as mediator, stratifying by ClinVar class and deleteriousness scores. PN resolves heterogeneity within the variant of uncertain significance (VUS) class beyond variant classification alone. In *GBA1* carriers with Parkinson’s disease, a mechanistically distinct, low-penetrance system, the same PN derived ordering of VUS holds, and PN is high despite modest penetrance. These results support causal necessity as a framework for genetic diagnosis, distinct from and complementary to variant classification, with implications for clinical and policy decisions in monogenic and common complex diseases.

## INTRODUCTION

Disease diagnosis operates in two distinct conceptual modes that are often entangled in clinical practice and decision making. Categorical diagnosis (*is-a-type-of* reasoning) is an annotation activity that labels the disease and determines its place in the medical coding ontology.^1–3^ A categorical diagnosis, like ischemic heart disease, may be secure while the drivers of the disease in a specific individual remain uncertain. In contrast, causal diagnosis^4^, which uses *is-a-cause-of* reasoning, considers a different question about the basis of disease, i.e. for which antecedents would the disease not have occurred if those antecedents had been absent.^5^ Causal reasoning can be crucial for management in many situations, particularly when reversing mechanistic drivers can directly halt, reverse, or at least slow the progression of phenotype.

In genetic diagnosis, the tension between the annotation and causal modes of reasoning is revealed in the formalism adopted for variant classification. In current practice, variant pathogenicity is a label assigned to variants based on evidence aggregated across cases and not intended as a contextual probabilistic statement about a given case or patient. According to Richards et al^6^ “Pathogenicity determination should be independent of interpreting the cause of disease in a given patient. For example, a variant should not be reported as pathogenic in one case and not pathogenic in another … Pathogenicity should be determined by the entire body of evidence in aggregate, including all cases studied, arriving at a single conclusion.” At present, the final diagnostic interpretation in individual patients depends on the clinical judgement of practitioners. The causal claim is implicit, and no counterfactual framework is available in the current standards and guidelines.

In contrast, causal reasoning is often at the heart of medical practice. To arrive at a causal diagnosis, physicians commonly work through the logic of possible models informally and intuitively.^7^ They narrow the scope of possibilities and arrive at the most likely mechanistic explanation as a guide to management. In genetic diagnosis, this means assessing whether genetic, polygenic, and/or environmental contributors are drivers of an individual’s disease. The increasing complexity and comprehensiveness of genetic testing together with the need to integrate genetic results with deep phenotyping – lab, imaging, physiologic observations and multi-omics profiles – challenge the intuitive heuristics of medical practice. The complexity of the available information for individual patients is juxtaposed with the ‘case-independent’ formalism of variant classification. We hypothesized counterfactual modeling might be useful to fill this gap and could provide a formalism to quantify the causal attribution of diagnosis as a probability that can be estimated from data according to the clinical and genetic profile a patient shares with a reference population.

Quantifying which antecedents are causally responsible for disease in a specific patient, requires methods for individual-level causal attribution. Pearl’s framework addresses this through probabilities of causation: Probability of Necessity (PN), Probability of Sufficiency (PS), and the Probability of Necessity and Sufficiency (PNS).^8^ These quantities involve counterfactuals – which represent the alternative world of individual outcomes when an exposure is absent. By definition these counterfactual states are unobservable at the individual level. In the context of germline genetic exposures, the counterfactual world corresponds to the life of an individual in the absence of a diagnostically relevant germline variant(s). For concreteness, one could imagine a monozygotic twin in which the variant in question have been edited out (although this idealization would still not match the counterfactual because it omits contingencies of development, epigenetic variation and postnatal exposures). Because individual level counterfactuals are unobservable, causal analysis focuses on counterfactual probabilities such as those in Pearl’s framework. Under certain assumptions, these counterfactual quantities are estimated from data and grounded in classical epidemiologic concepts. PN matches the epidemiologic concept of the etiologic fraction among the exposed, situating the consideration of genetic diagnosis within a longer tradition of attributable-risk reasoning.^9,10^ Dawid and Musio provide a comprehensive treatment of individual level causes which is deemed the Causes of Effects problem, clarifying the assumptions under which individual-level causal claims can be made.^11^ Gelman and Imbens^12^ caution that the determination of cause, the ‘‘why’’ question, requires multiple assumptions and has inherent limitations. Tian and Pearl^13^ formalize sharp bounds and point-identification results for PN and PNS under partial observability. More recent methods extend the framework to mediated pathways, enabling causal attribution through identified intermediates.^14–17^ Rubinstein et al. establish identification results for mediated probabilities of causation, including the total mediated probability of causation.^18^

Here we approach molecular diagnosis as a Causes of Effects problem. We focus on constitutional disease driven by germline variation; extensions to somatic variation are possible but outside the scope of this work. We frame molecular diagnosis as the probability that a DNA variant or ensemble of variants is necessary to cause a patient’s disease, such that, counterfactually, in the absence of those variant(s) the patient would not have had the disease (PN). Causal attribution under this definition is contingent on the patient’s presentation, selection for testing, the presence of alternative causes, and observable informative biomarkers. We show that the dependence on selection and alternative causes makes PN a property of a variant in a patient’s context rather than of the variant alone. We formulate the Probability of Mediated Necessity (PMN), which coincides with the total mediated probability of causation of Rubinstein et al.^18^, as the quantity that incorporates biomarker evidence into causal attribution. We explore the properties of both measures through simulation and by application to population-scale data. Our results support distinguishing these causal metrics from penetrance and from prior work on variant classification. Quantifying causal attribution reframes molecular diagnosis as a causal statement; crucially this statement is not a fixed determination. Instead, we advance causal understanding of molecular diagnosis as a probability, revisable as new evidence arrives, with consequences for diagnostic accuracy, therapeutic selection and patient outcome analyses.^19^

## METHODS

Pearl^8,13^ introduces three distinct forms of Probability of Causation - PN, PS, and PNS – and defines these as counterfactual quantities. In general, these quantities are bounds-identified rather than point-identified from experimental and/or observational data.^13^ Under the additional assumptions of *monotonicity* (a single direction of effect i.e. no variant is included in the analysis that is protective against disease in some individuals but is a driver of disease risk in others) and *exogeneity* (no unmeasured confounding), the bounds collapse to point estimates.^8^ We apply these concepts to models of genetic testing for diagnosis of a disease, noting that monotonicity, while highly plausible in genetics, is assumed rather than characterized at the individual variant level.

### Notation

Let *G* denote pathogenic variant carrier status (binary: 1 = carrier, 0 = non-carrier), *Y* denote disease outcome (binary), *M* denote mediator status (binary: 1 = present, 0 = absent), and *Z* denote the vector of measured covariates, which may include both confounders and alternative exposures. In the potential outcomes framework, each individual has a distinct outcome under each possible value of an exposure; for binary *G* these are *Y*(1) and *Y*(0), written in Rubin–Neyman notation as *Y*(*g*), the value *Y* would take under an intervention setting *G* = *g*. Both the conditional distribution of potential outcomes and the propensity for exposure may depend on *Z*.

The base model is *G* → *Y* (Figure 1A) indicating the variant *G* is a cause of disease *Y*. When multiple causes or a chain of causes are modeled, we define the compound potential outcome *Y*(*g, x*), the value *Y* would take under a joint intervention setting *G = g* and *X = x*. When *M* mediates the effect of *G*, *M*(*g*) denotes the value the mediator would take under *G = g*, and the natural-effect potential outcome is given by *Y*(*g*, *M*(*g)*). This notation also permits cross-world potential outcomes, in which *G* is set to *g* while M is held at a value distinct from *M*(*g*).

**Figure 1.**
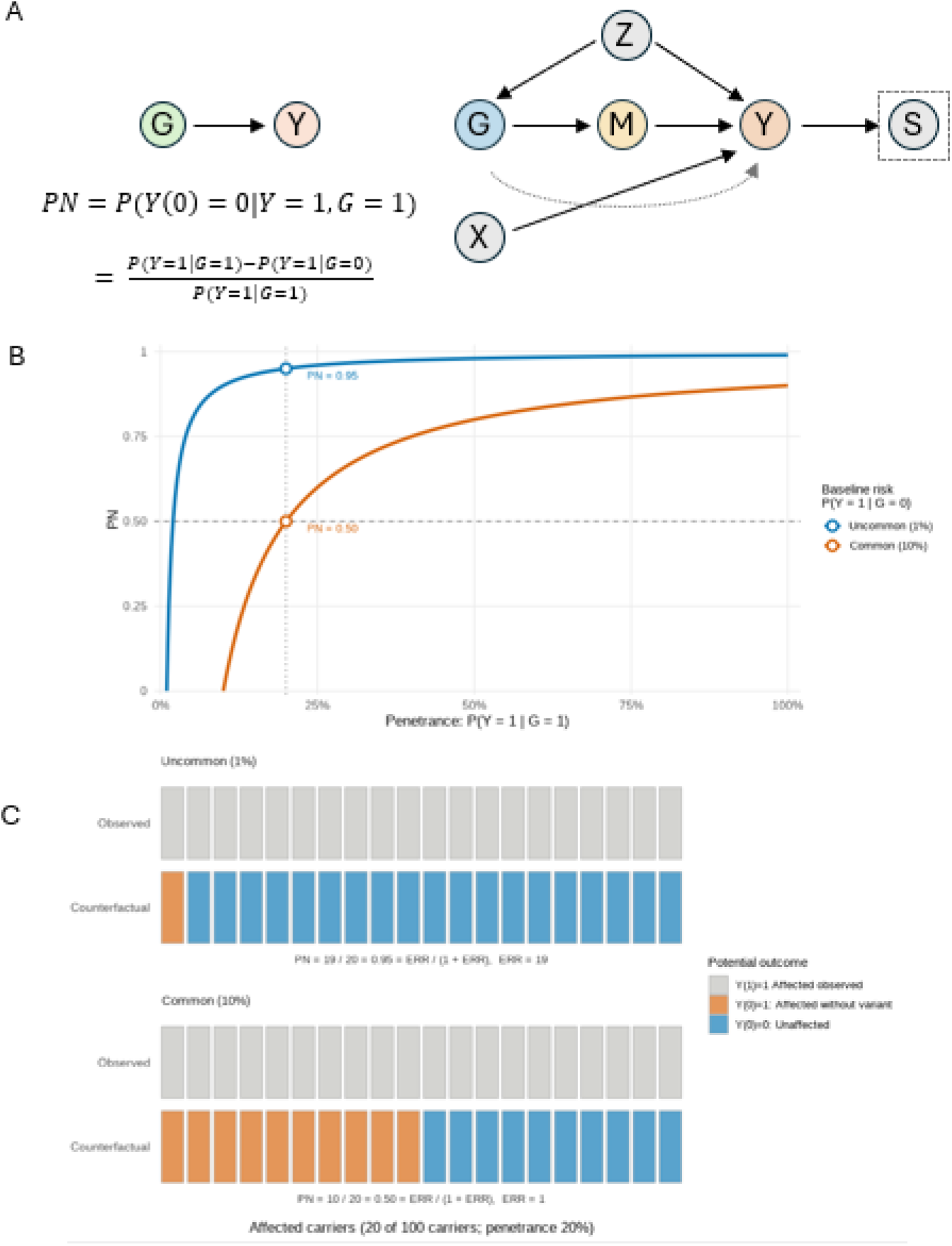
**A** Directed Acyclic Graph (DAG) Model of Genetic Effect on Disease. Nodes represent variables considered in this study and directed edges represent causal effects. A genetic variant (**G**) causes biochemical or structural perturbations which act as mediators (**M**) for the final effect on disease outcomes (**Y**). Confounding of G with Y could result in non-causal association of G and Y driven by common cause variables as represented by **Z**. Disease may result from other independent causes (**X**). Individuals may be recruited into the observational dataset based on their phenotype/disease thus acting as a selection variable (**S**). **B**. Dissociation of PN and Penetrance. Two disease populations were simulated in which the baseline risk of disease was 1% (uncommon) and 10% (common). Assuming only 20% penetrance, highlighted at the gray dotted line (P(Y=1|G=1)), PN was 0.95 for the uncommon disease scenario but only 0.5 in the common disease scenario. **C**. The underlying causal type distribution explains the difference in the two disease scenarios, where there are many fewer individuals with the Always (Y(0)=Y(1)=1) type when the disease is less frequent. The plot emphasizes the counterfactual structure that underlies the observed events.

We reserve ’phenocopy’ for its classical definition^20^ (environmentally-induced similar phenotype), distinguishing it from locus heterogeneity and other genetic architectures that can also produce disease independent of *G*. We use the Always-type fraction (π_A, vide infra) for the proportion of individuals who develop disease regardless of *G*, encompassing phenocopies, locus heterogeneity, and polygenic causation.

### Probability of Necessity (PN)

The primary estimand for this study is the Probability of Necessity (PN) which is defined counterfactually in the model *G* → *Y* (Figure 1A)

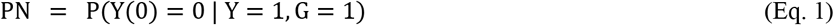

This is the probability that, among variant carriers who develop disease, disease would not have occurred in the absence of the variant. While conditioning on *Y*(1) = 1 rather than the observed *Y* = 1 would be the strict potential outcomes form, under consistency (Identification Assumption A1, vide infra) the two are equivalent. Under consistency (A1) and monotonicity (A3), PN is identified by the two-term formula (Pearl 2009, Eq. 9.31):

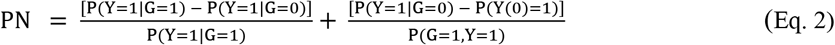

The first term is the excess risk ratio (ERR); under exogeneity and monotonicity the second term vanishes and PN equates to ERR. This quantity corresponds to the attributable fraction among the exposed. The penetrance P(*Y*=1|*G*=1) of the variant appears in both the numerator and the denominator of this term, but the PN requires more information than penetrance. The second term is the confounding correction. It requires the marginal counterfactual quantity P(*Y*(0)=1), the probability of disease that would be observed if the entire population were set to non-carrier status. This is distinct from the observed risk among non-carriers, P(*Y*=1|*G*=0), which is a conditional probability on the subpopulation that does not carry the variant. The two quantities coincide only when carrier status is unconfounded with the counterfactual outcome, and the second term measures their divergence. Its denominator is the joint probability P(*G*=1, *Y*=1), not the conditional P(*Y*=1|*G*=1).

Under monotonicity (no variants with an opposing effect) and exogeneity (A2: *Y*(0) ⊥ *G* | *Z*), the first term is sufficient to identify the PN. Under these assumptions, if there is confounding from factors *Z*, the causal quantity P(*Y*(0)=1) remains identified using the covariate adjustment formula:

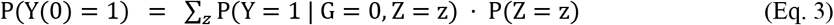

so that P(*Y*=1|*G*=0) = P(*Y*(0)=1). In this case we define PN for each level of Z-conditioning estimand:

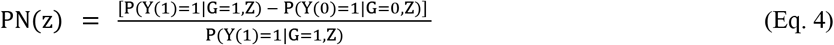

which, under A1–A3, is identified from observed data as:

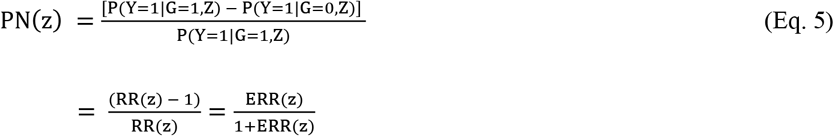

where RR = P(*Y*=1|*G*=1,*Z*) / P(*Y*=1|*G*=0,*Z*) is the covariate-adjusted risk ratio and ERR(z) = RR(z) – 1 is the excess relative risk. The marginal PN is obtained by averaging PN(z) over the covariate distribution of variant carriers with disease (see Statistical Models below).

For simplicity and exposition we implemented PN estimation using logistic regression models to estimate the conditional probabilities *P*(*Y* = 1 ∣ *G* = *g*, *Z*) from data and to parameterize simulation studies. For each analysis, we fit a single logistic regression model:

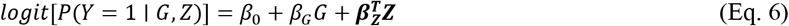

From this model, we obtained predicted probabilities *p̂*_1_ = *P*(*Y* = 1 ∣ *G* = 1, *Z*) and *p̂*_0_ = *P* (*Y* = 1 ∣ *G* = 0, *Z*) by setting *G* to 1 and 0 respectively while holding covariates *Z* constant. The PN was then calculated as 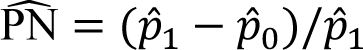, which can also be expressed as 1 − *p̂*_0_/*p̂*_1_ or equivalently 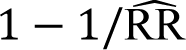
where 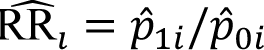 is the individual-level estimated risk ratio, which varies across covariate values under the logistic model. Individual-level PN estimates were computed for each carrier case using their observed covariate values, and the reported PN is the mean across carrier cases.

### Probability of Mediated Necessity (PMN)

Because PN varies with the strength of competing causes as well as prevalence; a biomarker that selectively marks individuals driven by the suspected genetic mechanism provides a way to improve inference for cases because the additional information can suggest a variant is functionally active in an individual patient. We extend the PN framework to quantify whether a genetic variant caused disease in the presence of observed biomarker information, and we term the resulting estimand the Probability of Mediated Necessity (PMN). Consider the causal structure *G* → *M* → *Y* (Figure 1A) with *G* additionally permitted a direct edge to *Y*. PMN is the probability that disease would not have occurred in the absence of the putative genetic mechanism i.e. *G* set to 0 and the mediator left at the value *M*(0), which is the potential outcome it would naturally take under *G* = 0; we condition on sampling among carriers who developed disease and who carry the biomarker abnormality:

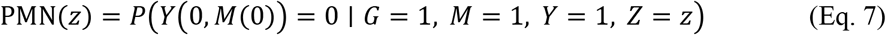

where Z are covariates. Conditioning on *M* = 1 restricts attention to cases with biomarker evidence that the mechanism was “on.” It enriches the stratum for individuals in whom the *G* → *M* → *Y* chain was plausibly active, without asserting that *G* caused the biomarker abnormality in any particular individual.

This estimand is the total mediated probability of causation δ̂(*x*) of Rubinstein, Cuellar and Malinsky (RCM).^18^ We retain the terminology of Pearl’s necessity/sufficiency hierarchy because of the parallel with PN and because it preserves the distinction from sufficiency. We write *δ*(*z*) for clarity when covariates *z* are also observed. By the composition axiom, *Y*(0, *M*(0)) ≡ *Y*(0) because intervening to set the mediator to the value it would have taken naturally for G=0 is not an intervention. PMN is therefore PN evaluated on *M* = 1 strata.

RCM (Theorem 1, eq. 12) identifies *δ*(*z*) from the observed-data distribution under assumptions A1–A9 below. Where *γ_g_*(*z*) = *P*(*M* = 1 ∣ *G* = *g*, *Z* = *z*), we can define the stratum weights

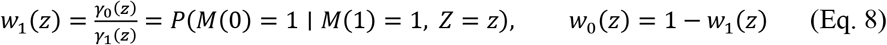

the second equality holds because of mediator monotonicity (A8a) and Bayes’ rule, with RCM’s Proposition 1 applied with *M* in place of *Y*. Then

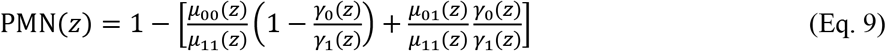

The numerator of Eq. 9 is *E*[*Y*(0, *M*(0)) ∣ *G* = 1, *M* = 1, *Y* = 1, *Z* = *z*], obtained by marginalising the counterfactual risk *P*(*Y*(0, *m*) = 1 ∣ *Z* = *z*) over the distribution of *M*(0) within the conditioning stratum (over *M*(0) ∣ *M*(1) = 1) rather than over the population distribution of *M*(0). The two ingredient counterfactual quantities are recovered from observables as

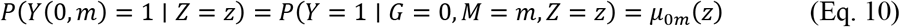

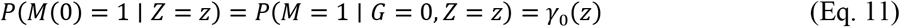

under assumptions A2, A4 and A5, and are then reweighted by Eq. 9 under A7 and A8.

An alternative construction that uses parallels to the mediation-formula literature, weights the two counterfactual risk arms by the marginal mediator distribution under non-exposure, (1 − *γ*_0_, *γ*_0_), rather than by (*w*_0_, *w*_1_):

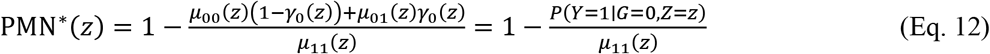

PMN^∗^ is an excess risk ratio comparing (*G* = 1, *M* = 1) against the entire non-carrier population. It differs from PN only in its denominator, and its numerator does not depend on the mediator at all. *^†^* Because *γ*_0_ ≤ *γ*_1_ implies *w*_1_ ≥ *γ*_0_, and because *μ*_01_ ≥ *μ*_00_ whenever the biomarker is a positive risk factor among non-carriers, the numerator of Eq. 9 is never smaller than that of Eq. 12. Hence

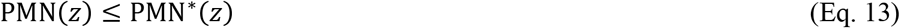

with equality only in the degenerate cases *γ*_1_ = 1 or *μ*_01_ = *μ*_00_. PMN^∗^ is therefore anti-conservative relative to the RCM-identified estimand: it overstates mediated necessity by exactly the factor

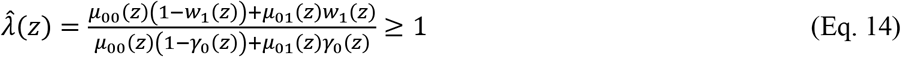

We caution that although λ̂ measures the cost of assuming *M*(0) ⊥ *M*(1), it is uninformative about violations of cross-world ignorability of *M* and *Y* (A7).

The same identification yields a decomposition of PMN into the necessity running through the mediator and the necessity not running through it (RCM eqs. 11 and 13):

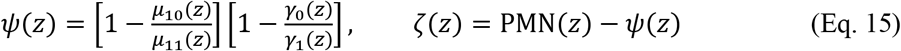

Here *ψ* is the probability that the mediating event occurred because of the variant and the outcome occurred because of the mediating event (indirect causation) and *ζ* is the residual attributable to pathways not through *M*.

Under parametric modelling assumptions for binary *Y* and *M*, PMN(*z*) requires two logistic regression models:

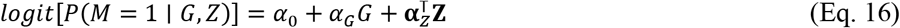

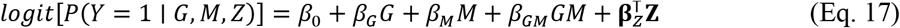

The four risk surfaces μ̂*_gm_*(*z*) and the two mediator probabilities γ̂*_g_*(*z*) are obtained as fitted values from Eqs. 16 and 17 evaluated at the covariate values of each carrier case, substituted into Eq. 9, and averaged over the (*G* = 1, *M* = 1, *Y* = 1) stratum. PMN^∗^uses the identical fits and differs only in the numerator weights. The *G* × *M* interaction in Eq. 17 is required: Eq. 9 evaluates the outcome model at all four (*g*, *m*) cells, and a main-effects model would impose a common odds ratio across them.*^‡^* We note that the max{0,⋅} operator biases lower bounds upward in finite samples.

### Identification Assumptions

Identification means that the targeted estimand can be expressed uniquely as a functional of the observed-data distribution. Identification of PN requires A1–A3; identification of PMN requires the additional assumptions A4–A9. A formal correspondence with the assumption set of RCM, together with the testability and consequence of violation for each, is given in Supplemental Table S1.

A1. Consistency. The observed outcome equals the potential outcome under the realized exposure and mediator: *M* = *M*(*g*) when *G* = *g*, and *Y* = *Y*(*g*, *m*) when *G* = *g* and *M* = *m*. For binary *G* this is the consistency equation *Y* = *G Y*(1) + (1 − *G*) *Y*(0). The assumption also precludes interference between individuals. [RCM Assumption 6.]

A2. Exogeneity (no unmeasured *G*–*Y* confounding). *Y*(0) ⊥ *G* ∣ *Z*. Conditional on measured covariates *Z*, the counterfactual outcome is independent of carrier status. Germline variant status is randomised at meiosis with respect to non-genetic exposures. Ancestry principal components in *Z* partially address population stratification.

A3. Outcome monotonicity in *G*. *Y*(1) ≥ *Y*(0) for all individuals: no carrier has lower disease risk as a consequence of carrying the variant. This is biologically defensible for most loss-of-function and deleterious variants given convergent functional, clinical and epidemiological evidence, though deleteriousness itself carries uncertainty. Monotonicity is untestable at the individual level; under violation, Pearl’s PN formula returns an upper bound.

A4. No unmeasured *G*–*M* confounding. *M*(*g*) ⊥ *G* ∣ *Z*. Carrier status is not confounded with mediator status beyond what is captured in *Z*. Together with A2 this is RCM’s joint randomisation condition *G* ⊥ {*Y*(1,1), *Y*(0, *m*), *M*(1), *M*(0)} ∣ *Z*. [RCM Assumption 8’.]

A5. No unmeasured *M*–*Y* confounding. *Y*(0, *m*) ⊥ *M* ∣ *G*, *Z*. The mediator–outcome relationship is unconfounded conditional on genotype and measured covariates. This is the most demanding of the single-world assumptions. Sensitivity of PMN to violations of A5 is addressed in the Supplementary Material.

A6. No unmeasured *G*-induced *M*–*Y* confounding. There exists no unmeasured *L* with *G* → *L*, *L* → *Y* and *L* → *M*, where *L* is not on the causal path from *M* to *Y*. Such an *L* would arise if *G* had pleiotropic effects on a variable that both influences *M* and reaches *Y* by a route other than *M*. A6 is not separately numbered by RCM; it is a substantive restriction on the causal graph under which A4, A5 and A7 are jointly credible.

A7. Cross-world ignorability. {*M*(0), *M*(1)} ⊥ *Y*(0, *m*) ∣ *Z* for *m* = 0,1. The mediator status an individual would exhibit, in either world, is independent given *Z* of the outcome that individual would experience in the absence of the variant. [RCM^18^ Assumption 9’.] Because the conditioning event {*G* = 1, *M* = 1} is, by consistency, the event {*G* = 1, *M*(1) = 1}, the estimand depends on the joint distribution of potential outcomes drawn from two distinct hypothetical worlds. No observed-data distribution constrains that joint law, so PMN is not point-identified without A7, and the assumption is untestable in principle. It is distinct from A5: A5 concerns confounding of the *M*–*Y* relationship within a single world, whereas A7 links quantities across worlds and cannot be addressed by measurement or adjustment.

The germline setting removes several mechanisms that threaten cross-world assumptions for modifiable exposures. Because *G* is fixed at conception and precedes all downstream biological processes, there is no time-varying confounding, no feedback from biomarker to exposure, and no selective exposure conditional on early outcomes. Individuals for whom *M*(0) = 1 carry that liability through polygenic and environmental background. Conditioning on *M* = 1 therefore enriches the analytic stratum for individuals at elevated counterfactual risk and A7 fails to the extent that such enrichment exceeds what the RCM weights (*w*_0_, *w*_1_) already accommodate. We emphasize that λ̂ estimated under A1–A9 quantifies the stratum enrichment that mediator monotonicity permits but it does not detect residual A7 violation.

A8. Mediator monotonicity in *G*, and outcome monotonicity in (*G*, *M*). Two distinct monotonicity conditions are required beyond A3,

A8a. *M*(1) ≥ *M*(0) for all individuals: no carrier has a lower biomarker value as a consequence of carrying the variant. This is the condition *P*(*M*(0) = 1 ∣ *M*(1) = 1) = *γ*_0_/*γ*_1_ (Eq. 9).

A8b. *Y*(1,1) ≥ *Y*(0, *m*) for *m* = 0,1: an individual who would not develop disease as a carrier with the biomarker present would not develop it as a non-carrier at either mediator level. Together these constitute RCM Assumption 7’. Both rest on the same variant-level aggregation of functional and computational evidence as A3, and both are untestable at the individual level.

A9. Positivity. *P*(*G* = *g*, *M* = *m* ∣ *Z* = *z*) ≥ *ε* > 0 for all (*g*, *m*) and all *z* in the support of *Z*, and *P*(*Y* = 1 ∣ *G* = 1, *M* = 1, *Z* = *z*) ≥ *ε* > 0. The first condition ensures that all four risk surfaces *μ_gm_*(*z*) are estimable; the second that the denominator of Eq. 9 is bounded away from zero. Positivity is the binding practical constraint in rare-variant applications, where the (*G* = 1, *M* = 1) cell may be sparse; it is assessed empirically.

### Simulation by Causal Type Composition

Our simulations employ a “TopDown” parameterization in which the population is specified as a mixture of latent causal types, and all observable epidemiological quantities are derived from this mixture. This method guarantees that the ground-truth (oracle) PN is known exactly, thereby providing an unambiguous benchmark against which estimators and estimates are evaluated.

Under the base model *G → Y*, each individual belongs to one of three latent response types defined by the pair of potential outcomes {*Y*(0), *Y*(1)}: Necessary (*Y*(0) = 0, *Y*(1) = 1, disease occurs only in the presence of the variant); Always (*Y*(0) = *Y*(1) = 1, disease occurs regardless of variant, representing phenocopies and other genetic causes); and Never (*Y*(0) = *Y*(1) = 0, disease does not occur regardless of variant) (Table S2). A fourth type is logically possible under an unrestricted model (the preventer (*Y*(0) = 1, *Y*(1) = 0), for whom the variant is protective) but this type is excluded by the monotonicity assumption *Y*(1) ≥ *Y*(0), imposed for all individuals such that P(*Y*(0)=1, *Y*(1)=0) = 0. In the germline genetics setting, monotonicity is justified on mechanistic grounds. A pathogenic allele that increases disease liability through a defined molecular mechanism is not expected to be protective in a subset of carriers, so the preventer type is assigned zero mass. Enforcing monotonicity as a construction constraint ensures that the simulated population contains no preventers of the assumed causal direction, and that the oracle PN computed from counted potential outcomes coincides exactly with the identification-formula PN.

Type proportions π_N, π_A, and π_I (with π_N + π_A + π_I = 1) determine all observable risks:

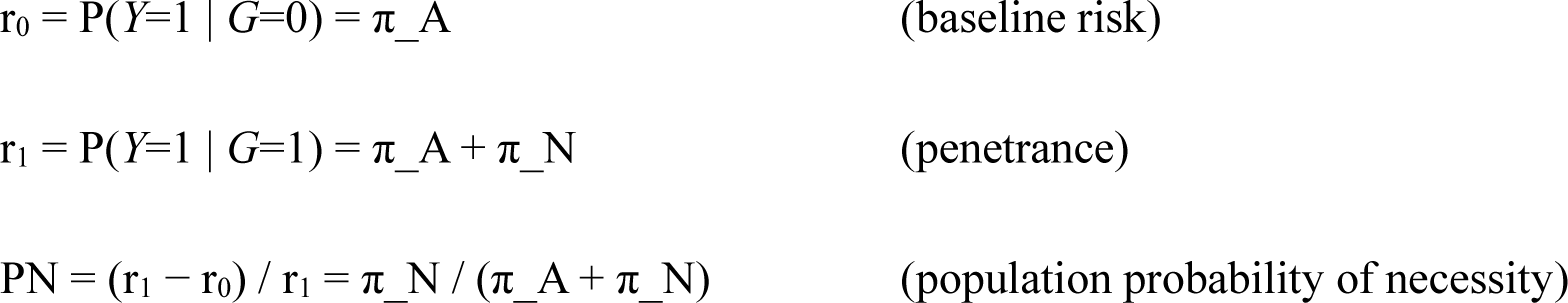

The oracle individual-level necessity PN_i_ takes the value 1 for individuals with *Y*(1) = 1 and *Y*(0) = 0 i.e. the Necessary-type carriers whose variant was counterfactually required for their disease, and 0 otherwise. The population PN is its expectation among affected carriers.

Unless otherwise noted, Monte Carlo scenarios simulated N = 10^6^ individuals per replicate with variant frequency P(*G*=1) = 0.002 and 50-100 replicates per swept parameter value, with potential outcomes assigned directly from the latent type structure so that the oracle PN equals the identification-formula PN up to sampling variation. Key quantiles of the sampling distribution of the PN estimator were derived from the 2.5^th^ and 97.5^th^ percentiles across Monte Carlo replicates. Scenarios that extend the base graph, adding an independent alternative cause (*G → Y ← X*), a selection node (*G → Y → S*), or a mediator (*G → M → Y*), are described in the corresponding sections below. The biomarker-mediated analysis extends the type structure from three to twelve latent types and is detailed separately.

### Penetrance–PN dissociation

Applying the base TopDown framework assuming monotonicity and exogeneity, we derived PN analytically across the full penetrance range under two baseline-risk scenarios, fixing π_A at 1% and 10%, corresponding to uncommon and common disease, respectively. For each scenario, π_N was swept from near zero to its maximum feasible value (1 − π_A), with π_I absorbing the remainder, tracing the full range of achievable (penetrance, PN) pairs. Because all quantities are well defined functions of the type proportions, no Monte Carlo simulation was required.

### Effect of Patient Selection

Selection of cases in a clinic-based study or other forms of sample selection could affect epidemiological measures of variant–disease association and consequently impact PN estimates. We extended the base framework with a selection node under the structure *G → Y → S*, where genotype influences disease status, which in turn determines selection probability *S* into the testing cohort. Selection was parameterized by λ = q₁/q₀, the ratio of selection probability for affected individuals (q₁ = P(*S*=1 | *Y*=1)) to unaffected individuals (q₀ = P(*S*=1 | *Y*=0)), with higher λ representing more aggressive case-based ascertainment. q₀ reflects referral driven by clinical suspicion, family history, or cascade testing in individuals who do not have the disease under study.

Each individual was assigned a causal type with scenario-specific probabilities, and potential outcomes were generated by shared-uniform coupling: a single draw U ∼ Uniform(0,1) per individual, with *Y*(0) = 1{U < p_Y0} and *Y*(1) = 1{U < p_Y1} for the type-specific risks p_Y0 ≤ p_Y1. This construction enforces monotonicity and guarantees that Always types satisfy *Y*(0) = *Y*(1) exactly, so that the causal-type label and the realized potential outcomes are consistent. Genotype was assigned independently with scenario-specific carrier frequency and observed disease status was *Y* = *Y*(G).

Three disease scenarios were examined spanning a range of penetrance and baseline rate: Scenario A (80% Necessary, 2% Always, 18% Never; penetrance 12.8%, baseline risk 1.6%, oracle PN 87.6%); Scenario B (60% Necessary, 15% Always, 25% Never; penetrance 53.8%, baseline 14.8%, oracle PN 72.6%); Scenario C (50% Necessary, 30% Always, 20% Never; penetrance 61.0%, baseline 26.0%, oracle PN 57.4%). Selection enrichment λ was swept across {1, 2, 5, 10, 20} with q₀ fixed at 0.05. We compared three quantities in the simulated populations. The oracle is PN_cf_ = P(*Y*(0) = 0 | *G*=1, *Y*=1), was evaluated directly from the simulated potential-outcome vectors of the unselected population. Because genotype is assigned independently of type and of U, this equals P(*Y*(0)=0 | *Y*(1)=1) and is computed over all simulated individuals. The naive estimate is the excess-risk ratio (r̂₁ − r̂₀)/r̂₁ computed in the selected sample. Finally, the selection-adjusted estimate applies a closed-form inversion of the selected-sample risks, r = r_sel·q₀ / (q₁ − r_sel(q₁ − q₀)), before forming the same ratio. This expression depends on q₁ and q₀ only through λ, so knowledge of λ alone is sufficient for correction. The adjustment assumes *S* ⊥ *G* | *Y*, that selection depends only on disease status and not directly on genotype. Causal-type composition was tracked under two denominators: among affected carriers (*G*=1, *Y*=1, *S*=1) and among all selected carriers (*G*=1, *S*=1), with counts pooled across replicates before forming proportions (Figure S1). Estimates were computed across 100 Monte Carlo replications, with the population regenerated per replicate; shaded bands report 2.5th–97.5th percentiles across replicates.

### Modeling Alternative Causes

In many diseases, alternative independent causes compete with the variant of interest to drive disease in the affected population. These include monogenic locus heterogeneity, polygenic effects, and non-genetic factors. We modelled the alternative cause as a standardized polygenic score *X* ∼ N(0,1), independent of *G* by construction (*X* ⊥ *G*). This instantiates alternative causation under the DAG *G → Y ← X*.

To isolate the compositional mechanism underlying PN heterogeneity in the presence of competing causes, we used fixed parameters in the TopDown causal type framework. The purpose is to evaluate the GLM estimator against a closed-form PN at a single configuration where effect magnitude and type composition are known exactly. Carrier status and polygenic score were drawn independently: *G* ∼ Bernoulli(0.005), so that 0.5% of individuals carry the variant, and X ∼ N(0,1). Each individual was assigned one of three latent causal types by multinomial draw: Necessary (40%), Always (20%), Never (40%). An indicator of *X*-driven disease was drawn as A ∼ Bernoulli(*expit*(α + β_X_X)), with α = logit(0.10) and β_X = log(1.8), giving approximately 10% background probability at *X* = 0 and an odds ratio of 1.8 per standard deviation. Potential outcomes were assigned as follows. *Y*(0) = 1 if type is Always or A = 1, and 0 otherwise. *Y*(1) = 1 if type is Always, type is Necessary, or A = 1, and 0 otherwise. These conditions enforce monotonicity *Y*(1) ≥ *Y*(0). The type label does not by itself fix the realized potential outcome: a Necessary-type individual with A = 1 has Y(0) = Y(1) = 1 and contributes no necessity. Overall outcome prevalence was 0.291 modeling a common complex trait.

Four quantities were compared within each stratum of *X*. Analytic PN is the closed-form value implied by the type mixture and the background-risk logistic. Because the mixture is linear in p(x) = P(A = 1 | *x*) on the probability scale, P(*Y*(0) = 1 | *x*) = π_A + (1 − π_A)·p(*x*) and P(*Y*(1) = 1 | *x*) = (π_A + π_N) + (1 − π_A − π_N)·p(*x*), where π_A and π_N are the Always and Necessary proportions. Analytic PN is the corresponding excess-risk ratio, evaluated by averaging each component over the observed *X* within a stratum. It carries no Monte Carlo error and serves as the ground-truth reference. Oracle PN is the mean of the indicator 1{Y(1) = 1, Y(0) = 0} among affected carriers (G = 1, Y = 1), taken from the simulated potential-outcome vectors. It is the finite-sample realization of the analytic quantity and carries binomial variability that is substantial where carriers are sparse. Empirical PN is the stratum-specific plug-in (p̂₁ − p̂₀)/p̂₁, where p̂_g_ is the observed proportion with Y = 1 among individuals with G = g in that stratum. GLM PN uses a logistic interaction model, *logit* P(*Y* | *G, X*) = α + β_G_*G* + β_X_*X* + β_GX_*GX*, fitted once on the full sample. An additive model omitting the interaction term was also fitted for comparison of β_G_. Within each stratum, fitted risks were obtained by averaging individual-level predictions over the observed *X* values in that stratum under *G* = 0 and *G* = 1, and PN was formed as (p̄₁ − p̄₀)/p̄₁. This is a ratio of stratum-averaged risks (not a stratum average of individual-level PN).

*X* was divided into 20 fixed-width bins of 0.5 SD spanning [−5, 5]. Bins containing fewer than 50 carriers were excluded, retaining 10 bins with midpoints from −2.25 to 2.25 SD. No estimate or bootstrap replicate fell outside [0,1], so no truncation was applied. Each bin is indexed by its midpoint *x*, and stratum-specific quantities are written as functions of it, e.g. PN(*x*). Uncertainty was quantified by bootstrap, with 200 replicates and percentile intervals at the 2.5th and 97.5th quantiles. The two estimators require different resampling units. Empirical PN is a stratum-specific statistic, so its bootstrap resamples individuals with replacement within the stratum and recomputes the plug-in. GLM PN is a full-sample statistic, since the point estimate derives from a single fit to all individuals. Its bootstrap resamples the full sample, refits the interaction model once per replicate, and predicts into every stratum at the observed *X* values. Refitting within a single 0.5 SD stratum was rejected: over that span *X* is close to constant, the resamples are frequently quasi-separated, and the interaction coefficient diverges without triggering a convergence failure. All 200 full-sample refits converged. Coverage was assessed against analytic PN. The oracle is reported without an interval. These intervals quantify sampling variability only. They do not reflect identification uncertainty, which is governed by monotonicity and exogeneity (A2, A3).

### Simulation of Biomarker-Stratified Causal Attribution

Biomarkers can be causal intermediates and when they are they give information that the DNA variant causal path is active. We used simulation to investigate some of the effects and limitations of using biomarker measurements in the PN framework. This framework facilitates evaluation of how conditioning on a mechanistic biomarker (*M*) affects PN and PMN for a genetic variant (*G*) and disease (*Y*) under the causal structure *G → M → Y*. This biomarker analysis extends the TopDown simulation framework from three to twelve latent causal types (Table S8) to accommodate the mediator structure. This modeling framework uses a coupling table P(*U*_Y | *U*_M), which specifies the conditional distribution of outcome response types given mediator response types, making the joint potential-outcome structure explicit and ensuring coherence of the probabilities. These types derive from the 64 possible combinations of *G, M*, and *Y* potential outcomes, restricted by the monotonicity assumption, and then further restricted by an assumption of no cross-world effects. Causal types (Table S8) arise from three mediator types - M-Never (*M* = 1 only at a low background rate regardless of *G*), M-Caused (*M* = 1 at high rate when *G* = 1, reverts to low rate under *G* = 0), and M-Always (*M* = 1 regardless of *G*) which are summarized in Table S7. These three M-types are crossed with four outcome types: Y-Never, Y-M-only (disease driven by biomarker elevation), Y-G-only (disease driven by variant via a direct pathway), and Y-Always. The logical constraint P(Y-M-only | M-Never) = 0 is enforced by setting the corresponding coupling table cell to zero (mediator coherence). Oracle values of PN and PMN were computed directly from assigned potential outcomes; GLM-based estimates were obtained using logistic regression models for P(*M* | *G*) and P(*Y* | *G*, *M*), with counterfactual risks computed by marginalizing individual-level predictions over the *G* = 0 mediator distribution.

Four sweep analyses were conducted. Each point on each axis comprises 50 replicates of 10⁶ individuals at carrier frequency 0.002, with per-replicate seeds derived deterministically from the axis, the sweep index, and the replicate index. The biomarker specificity for G axis (Figure 4A) varied the rate of the biomarker positivity in the G=0 strata jointly across the M-Never and M-Caused mediator types over 2%, 5%, 10%, 20%, 30%, and 40%, with all other parameters fixed. The background disease burden axis (Figure 4B) varied the rate of disease arising from causes entirely outside the *G → M → Y* pathway from 0% to 30% in steps of 5%, with the Y-always mass in the M-Never row held fixed. Background risk enters as an independent competing cause applied identically to every (*G, M*) cell, P(*Y* = 1) = b + (1 − b)p. Here b is the probability of disease from causes acting outside the *G → M → Y* pathway and p is the cell-specific risk in the absence of those causes. The direct *G → Y* cause axis (Figure 4C) varied the mass of the Y-g-only causal type from 5% to 35% in steps of 5%. A Y-g-only individual has *Y*(1, *m*) = 1 and *Y*(0, *m*) = 0 for both values of *m*, so disease occurs if and only if the variant is present and the mediator is causally irrelevant. The mass is set to a common value in every U_M row.*^§^*Each row donates the added mass from its Y-never cell. Y-never and Y-g-only individuals are observationally identical whenever *G* = 0, so this rule leaves the joint distribution of *M* and *Y* among non-carriers unchanged and confines the perturbation to the carrier stratum. The cross-world dependence axis (Figure 4D) varied the dependence between mediator type and outcome type, which is the content of assumption (RCM 9′). The coupling table was interpolated between the independence table and the baseline table,

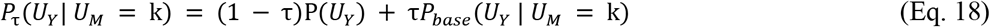

where P_base_ is the baseline coupling table, P(U_Y_) = Σ_k_ P(U_M_ = k)P_base_(U_Y_ | U_M_ = k) is its U_M_-weighted marginal, and τ is the weight placed on the baseline conditional distribution. The axis was swept over τ ∈ {−0.4, −0.2, 0, 0.2, 0.4, 0.6, 0.8, 1.0}. At τ = 0 every row equals the marginal, outcome type is independent of mediator type, and (RCM 9′) holds exactly. At τ = 1 the table is the baseline. Negative τ reverses the sign of the dependence relative to baseline. Because the interpolation is linear and the independence table is the U_M_-weighted marginal of the baseline table, the marginal distribution of outcome types is preserved at every τ, so total Y-always mass and total variant-causal mass are held fixed and cross-world dependence is the only quantity that moves. Mediator potential outcomes were drawn coupled at the individual level, so (RCM 7′) holds pointwise and the axis isolates (RCM 9′). Under coupled draws the counted counterfactual quantities PN_cf_ and PMN_cf_, rather than the individual-level averages PN_oracle_ and PMN_oracle_, are the targets against which both estimators are scored. The axis in Figure 4D is labelled by log RR = log[P(Y-always | M-always) / P(Y-always | M-never)], a signed projection of the table onto one cell pair.

### Empirical Study Population and Data Source

Data were drawn from the UK Biobank (UKB), a prospective population cohort of approximately 502,000 participants aged 40–69 years recruited in the United Kingdom between 2006 and 2010. Whole-exome sequencing (WES) data were accessed via the UKB Research Analysis Platform (RAP). Individual-level analyses were conducted exclusively within the RAP environment in compliance with UKB data access terms. UK Biobank holds Research Tissue Bank approval from the North West Multi-centre Research Ethics Committee (REC reference 21/NW/0157; IRAS project ID 299116), under which research conducted using data supplied by the resource does not require separate project-specific ethical approval. All UK Biobank participants provided written informed consent at recruitment, including consent for linkage to national health records. Participants who have withdrawn consent were excluded from all analyses in accordance with UK Biobank procedures. This research was conducted under UK Biobank Project ID 98786. No participant re-contact was undertaken and no new data were collected from participants. The analyses reported here used de-identified individual-level data supplied under the UK Biobank Material Transfer Agreement and were conducted exclusively within the UK Biobank Research Analysis Platform. The authors held no access to identifiers and no means of re-identifying participants. The Institutional Review Board of Baylor College of Medicine reviewed this activity and determined that the analysis is exempt under 45 CFR 46.104(d)(4).

The primary exposure of interest was heterozygous carrier status for a pathogenic or likely pathogenic *LDLR* variant (*G* = 1) versus non-carrier status (*G* = 0). Carriers were identified from the UKB WES release (500k exome, field 23157). The primary outcome (*Y*) was ischemic heart disease (IHD), defined by the presence of ICD-10 codes I20–I25 in hospital episode statistics or primary care records, or self-reported diagnosis at baseline assessment. The mediating variable (*M*) was diagnosis of hyperlipidemia (E78) prior to the first diagnosis date of IHD. Measured confounders (*Z*) comprised the first 4 genetic principal components (ancestry PCs), age at recruitment, and sex. The analytic sample comprised 1844 heterozygous LDLR variant carriers and 488260 non-carriers after exclusions (Table 1). Among carriers, 344 had a recorded IHD diagnosis of which 287 had a recorded E78 code.

**Table 1.** Counts of Cases and Controls – UKB Ischemic Heart Disease.

| Count (Heart Disease / LDLR ) |  |  |  |
| --- | --- | --- | --- |
|  | Y=0 | Y=1 | n(G) |
| G=0 | 430677 | 57583 | 488260 |
| G=1 | 1500 | 344 | 1844 |
| n(Y) | 432177 | 57927 | 490104 |
| Count (E78\|Y,G) |  |  |  |
|  | Y=0 | Y=1 | n(E78\|G) |
| G=0 | 90337 | 36894 | 127231 |
| G=1 | 697 | 287 | 984 |
| n(E78\|Y) | 91034 | 37181 | 128215 |

### *LDLR* Variant Computational Scoring and Quantile Stratification

To evaluate whether PN provides resolution within the VUS class we computed affected PN for *LDLR* variant carriers stratified jointly by ClinVar class and score tertile. Four computational deleteriousness scores were used, differing in the methods used to define the training labels. ESM-1b is a protein language model trained by self-supervision on sequence alone and uses no variant labels. AlphaMissense fine-tunes AlphaFold structural and evolutionary representations on weak labels derived from human and primate allele frequencies. CADD is trained to discriminate derived alleles from simulated de novo variants. REVEL is a random forest trained directly on curated disease mutations from HGMD, with component scores that are themselves supervised on curated disease sets. Only REVEL depends on clinically annotated labels during model fitting. AlphaMissense scores are monotonically recalibrated against a ClinVar evaluation set; because the analysis uses within-pool tertiles rather than published classification thresholds, this recalibration is rank-preserving and does not enter the stratification. Variants were assigned to tertiles of each score independently. Because individual-variant PN estimation is infeasible for singleton or near-singleton carriers, *G* was defined as carrier status within each ClinVar × tertile stratum, and individual-level affected PN scores were computed for affected carriers (*G*=1, *Y*=1) within each stratum using the marginal standardization formula above. Tertiles were chosen as a partition that preserved adequate cell size for stable bootstrap estimation; the number of bins was determined by examining CI width stability as a function of bin count, and three tertiles represented the point at which further subdivision produced materially wider confidence intervals without additional discriminative resolution. Affected PN values below zero, observed in benign variant strata, are reported unclamped; these reflect strata in which carrier IHD risk is at or below background and serve as a lower anchor for the scale. The same procedure was applied to *GBA1* variants with Parkinson’s disease as the outcome for cross-disease validation.

### Cross-Disease Validation

To evaluate whether the PN framework generalizes across gene–disease systems, we replicated the tier-stratified PN analysis in a second gene–disease pair: *GBA1* variants and Parkinson’s disease (PD). *GBA1* encodes glucocerebrosidase; heterozygous *GBA1* variants are a major genetic risk factor for PD, with a distinct biological mechanism (lysosomal dysfunction and α-synuclein aggregation) from the LDL-C–mediated pathway examined for *LDLR*. The same tier classification scheme (T1, T2, T2_disc) was applied to *GBA1* variants, and PN was estimated within each tier using the same logistic regression and bootstrap framework, with PD diagnosis (ICD-10 G20) as the outcome. ClinVar classification (pathogenic/likely pathogenic, VUS, benign/likely benign) was used as an independent label for validation.

### *LDLR* All Variant Classification and Functional Tier Assignment

To investigate whether PN/PMN could be applied comprehensively to all variants in an example disease gene, *LDLR* variants were classified into functional tiers based on predicted molecular consequence, independent computational pathogenicity scores, and gnomAD v4 allele frequency. Tier assignment was performed by a custom Python script applied to de-identified variant-level data. ClinVar classification (pathogenic/likely pathogenic, VUS, benign/likely benign) was retained as an independent cross-validation label and was not used in tier assignment.

Consequence tiers were assigned in the following priority order. T_splice_ (splice-disrupting) was applied as an override to any variant with a SpliceAI maximum delta score ≥ 0.50, regardless of VEP sequence ontology (SO) term. T1 (high-confidence loss-of-function) comprised variants with a LoF SO term (stop-gained, frameshift, canonical splice-site ±1/2, start-lost, stop-lost) and a coding VEP consequence confirmed by an SO allowlist. The coding VEP SO allowlist, requires the VEP-annotated consequence to belong to a pre-specified set of coding SO terms in addition to the SO-level LoF annotation. This prevents deep intronic indels with incidental LoF SO terms from being incorrectly assigned to LoF tiers. For missense and in-frame indel variants, a voting scheme was applied using three independent deleteriousness metrics: REVEL (strong vote: ≥ 0.75; moderate: 0.50–0.75), AlphaMissense used as a continuous deleteriousness score (strong: ≥ 0.80; moderate: 0.56–0.80), and ESM1b log-likelihood ratio (strong: ≤ −7.5; moderate: −7.5 to −5.0). CADD phred served as a moderate-only fallback (threshold: ≥ 20) applied only when all three primary scores were absent for a given variant. T2 (concordant strong missense) required ≥ 2 strong votes; T2_disc (discordant strong missense) had exactly 1 strong vote; T3 (moderate missense) had ≥ 1 moderate vote and 0 strong votes. All remaining variants, including synonymous, non-coding variants not meeting T_splice criteria, and missense variants with no qualifying votes, were assigned T4.

Each variant was additionally stratified by gnomAD v4 non-Finnish European (NFE) allele frequency: F1 (ultra-rare: AF < 0.01%; raw threshold 0.0001), F2 (rare: 0.01–0.1%), F3 (low-frequency: 0.1–1%), and EXCLUDE (common: AF ≥ 1%). Variants absent from gnomAD were assigned F1. The primary analysis was restricted to F1 variants. F2 and F3 strata are reported in sensitivity analyses. The final bin identifier concatenates consequence and frequency tiers (e.g., T2/F1).

Individuals carrying variants in more than one bin were assigned to a single analysis bin by a pre-specified priority order favoring higher-consequence tiers: T1/F1 > T2/F1 > T_splice_/F1 > T2_disc/F1 > T3/F1 > T4/F1, with AF sub-tiers ranked equivalently within each consequence tier. For PN estimation, the T3 and T4 bins were pooled into a single low-evidence stratum (T3T4l_ow_); this stratum serves as an internal negative control and is expected to have PN near zero.

### Statistical Models and Covariates

Conditional probabilities required for PN and PMN estimation were obtained by logistic regression with marginal standardization. Two binary outcome models were specified with identical linear predictor structure. The primary outcome model estimated P(*Y* = 1 | *G, Z*):

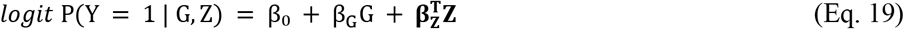

The joint outcome model estimated P(*Y_M_* = 1 | *G, Z*), where *Y_M_* = I(*Y* = 1) · I(*M* = 1) is the indicator of the composite event {IHD with E78 diagnosed before IHD}:

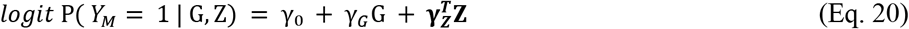

Both models included age, sex, a standardized CAD polygenic risk score (PRS), and the first four ancestry principal components as components of **Z**. Ancestry principal components address confounding by population structure. Age, sex, and PRS are not confounders of carrier status, which is independent of them; they enter as modifiers or proxies for competing causes of ischemic heart disease. Predicted risks are averaged over the observed covariate distribution of the working dataset before the ratio is formed, so the reported bin-level PN and PMN are marginal over polygenic background.

For each bin, a working dataset was constructed from carriers in that bin combined with a random subsample of non-carriers at a 100:1 ratio, drawn once prior to all analyses for that bin. Both models were fitted to this working dataset. Marginal standardization was applied by setting *G* = 1 or *G* = 0 for each individual while retaining observed covariates, then averaging predicted probabilities across the dataset:

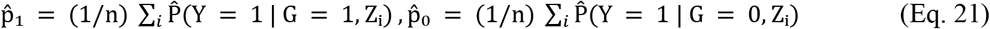

and analogously p̂₁ₘ and p̂₀ₘ from the joint outcome model. Bin-level PN and PMN were then computed as:

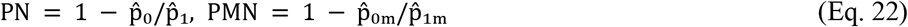

The composite outcome formulation for PMN is numerically equivalent to computing PN within the {*Y* = 1, *M* = 1} stratum when the direct *G → Y* effect is negligible (PMN identification formula under assumptions A1–A9). Due to complete separation in the logistic models for the T_splice_/F1 stratum (zero IHD events among T_splice_/F1 non-carriers in the relevant covariate strata), individual-level scoring was not performed for this tier but bin-level estimates are reported.

Bootstrap confidence intervals (95%) were computed using 1000 resamples. Because *LDLR* variant carriers are rare relative to non-carriers in UKB, a pre-thinning step was applied before bootstrapping: non-carriers were subsampled without replacement to a ratio of 100:1 (non-carriers to carriers) once prior to all bootstrap iterations, producing a pre-thinned dataset of manageable size that retains the asymptotic properties of the full-data estimate. Bootstrap resampling was then applied to this pre-thinned dataset with replacement, refitting both logistic models and recomputing PN and PMN on each resample. Confidence intervals were obtained from the 2.5th and 97.5th percentiles of the bootstrap distribution.

### Variant Holdout Validation

To evaluate tier-level PN estimates in unseen cases, a random 10% of IHD cases were withheld from model fitting prior to all analyses. Holdout IHD cases carrying an *LDLR* rare variant (n = 22) were scored by assigning the bin-level PN and PMN estimates from the training data according to each individual’s assigned bin. Holdout cases were additionally characterized by E78 status relative to IHD date, CAD PRS tertile, and ClinVar classification of the carried variant. Cases with E78 diagnosed before IHD received a PMN assignment in addition to PN; cases without antecedent E78 received PN only, with a note that the lipid pathway was not confirmed.

## RESULTS

### Probability of Necessity is Different from Penetrance

Penetrance is the primary metric by which clinical geneticists assess the causal weight of a variant. However, penetrance conflates two distinct populations of affected carriers: those for whom the variant was necessary to develop disease (Necessary-type), and those who would have developed disease regardless of variant status (Always-type, composed of phenocopies and those with independent genetic factors such as polygenic risk). PN quantifies the Necessary-type fraction among affected carriers and is therefore distinct from penetrance. It depends additionally on other factors including the baseline disease risk which influences both carriers and non-carriers.

To illustrate this dissociation, we derived PN analytically across the full range of penetrance values under two baseline risk scenarios: an uncommon disease (r0 = 1%) and a common disease (r0 = 10%). At 20% penetrance PN is 0.95 for the uncommon disease and 0.50 for the common disease (Figure 1B). Among 100 affected carriers at this penetrance, an average of 95 are Necessary-type in the uncommon disease versus an expected 50 in the common disease (Figure 1C). This difference is fully explained by the excess relative risk (ERR): ERR = 19 when r0 = 1% versus ERR = 1 when r0 = 10%, at identical penetrance. Because PN = ERR / (1 + ERR), a variant that appears weak by penetrance criteria can nonetheless be the necessary cause of disease in the great majority of affected carriers, provided the disease is sufficiently uncommon.

### Bias Introduced by Case Selection

Genetic testing is ordered by clinicians for patients based on clinical indications; therefore, ordering of a test can be viewed as a type of selection on the target population. We asked whether selection of patients for genetic testing based on clinical presentation introduces systematic bias into naive PN estimation, and whether this bias varies across disease contexts with different penetrance and alternative-cause rates. Selection is modeled as a binary variable (S=1 for tested individuals) and parameterized as λ = q₁/q₀, the ratio of selection probability for affected individuals (q₁ = P(*S*=1 | *Y*=1)) to unaffected individuals (q₀ = P(*S*=1 | *Y*=0)).

We adopted a ‘TopDown’ simulation strategy based on the potential outcomes framework. Rather than simulating genetic architecture and observing emergent biases, we explicitly specified latent causal types (Necessary, Always, Never) corresponding to potential outcomes *Y*(0) and *Y*(1). We simulated populations of 10⁶ individuals under three disease scenarios with contrasting penetrance and alternative-cause proportions, sweeping λ across {1, 2, 5, 10, 20} with q₀ fixed at 0.05. Each scenario was replicated 100 times. Naive PN estimates were systematically biased downward as ascertainment intensity increased, in all three scenarios (Figure 2). Under unbiased selection (λ = 1) the naive estimate recovered the oracle PN in every scenario. As λ increased the naive estimate progressively underestimated PN; at λ = 10 the shortfall was 10.9% in Scenario A, 41.5% in Scenario B, and 40.3% in Scenario C, and at λ = 20 the Scenario C estimate fell to 9.7% against an oracle of 57.4%.

**Figure 2.**
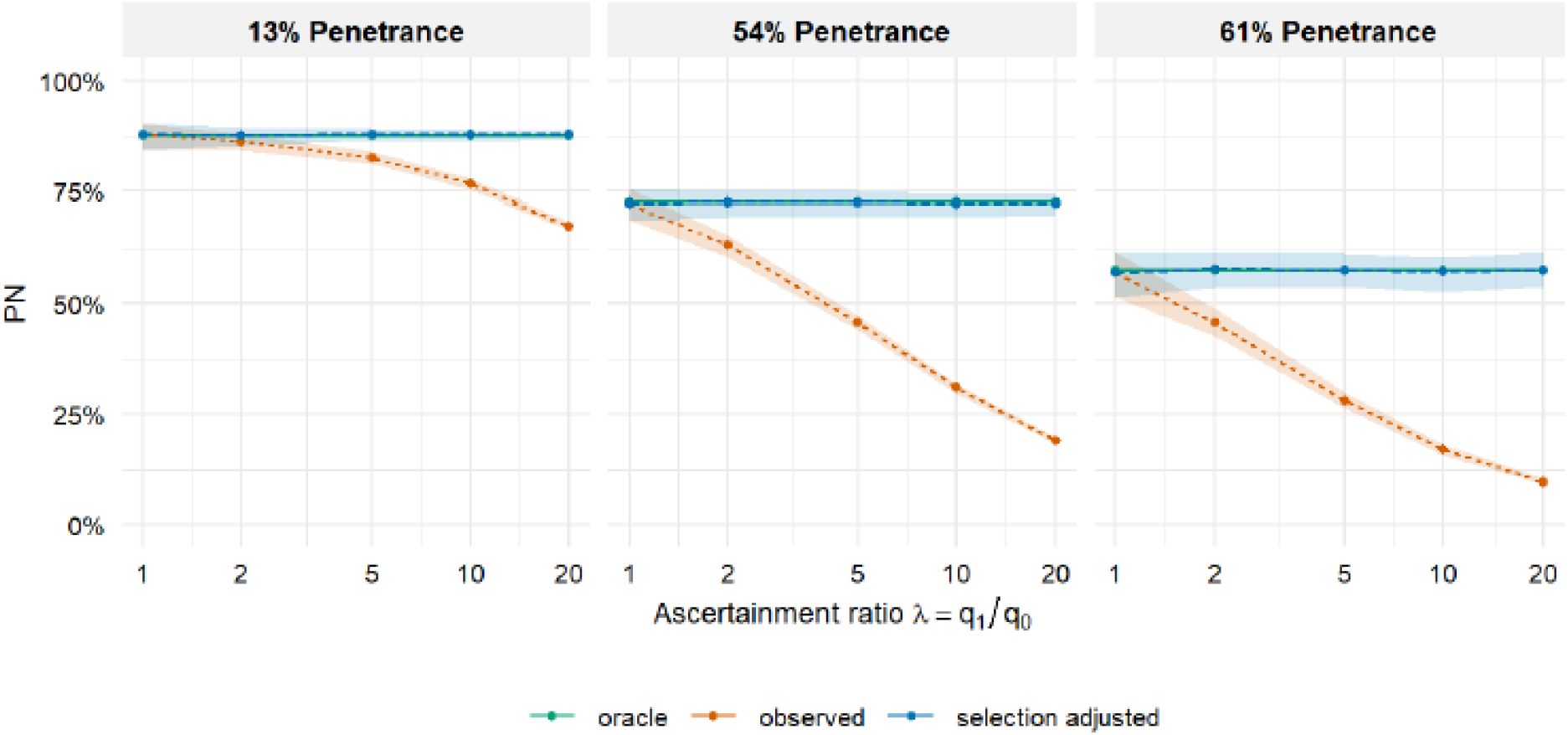
Ascertainment bias in PN estimation across three disease scenarios illustrating penetrance–PN dissociation. Each panel shows PN as a function of the ascertainment ratio λ = q₁/q₀, the likelihood ratio of referral considered as a test for disease. Scenarios: A (penetrance 13%, oracle PN 88%, baseline risk 1.6%); B (penetrance 54%, oracle PN 73%, baseline risk 14.8%); C (penetrance 61%, oracle PN 57%, baseline risk 26.0%). **Oracle** (green): PN_cf_ = P(Y(0)=0 | G=1, Y=1), evaluated directly from the simulated potential-outcome vectors of the full unselected population. **Observed (Naïve)** (orange): PN estimated from the ascertained subsample without correction, showing systematic downward bias that worsens with λ. **Selection adjusted** (blue): PN recovered by inverting the ascertainment process using known q₁ and q₀; recovery is exact under S ⊥ G | Y, but requires λ, which is identifiable only against an external population prevalence. Shaded bands are 2.5th–97.5th percentiles across 100 Monte Carlo replications (N = 10⁶ per replicate).

The precision of the naive estimate improved over the same range as its accuracy deteriorated: in Scenario C the interval half-width narrowed from 5.1 to 0.7 percentage points between λ = 1 and λ = 20, since more intense ascertainment yields more affected carriers. When the selection parameters are known, PN was recovered exactly. The selection-adjusted estimator returned the oracle PN to within 0.4 percentage points across all λ values in all scenarios (Figure 2, blue), confirming that the bias is correctable, in principle, under *S* ⊥ *G* | *Y*. Because the inversion depends on q₁ and q₀ only through their ratio, knowledge of λ alone is sufficient. The bias does not arise from a change in the causal-type composition of the case series. Among affected carriers, type composition was invariant in λ in all scenarios (Figure S1A); among all selected carriers, composition shifted only by depletion of Never types (Figure S1B). Instead, ascertainment drives both P(*Y*=1 | *G*=1, *S*=1) and P(*Y*=1 | *G*=0, *S*=1) toward unity, compressing the excess-risk ratio (r₁ − r₀)/r₁ from which PN is computed.

### Effect of Alternative Causes

The process of making a diagnosis requires consideration of alternative explanations for the presenting signs and symptoms. We modeled this by treating polygenic variation as an independent cause (Figure 1B, *G → Y ← X*). A population of N = 10⁶ individuals was simulated under the TopDown causal-type framework (40% Necessary, 20% Always, 40% Never). The variant *G* was introduced at carrier frequency 0.5%, and *X* ∼ N(0,1). These parameters yielded overall disease prevalence 0.291. Because carriers constitute 0.5% of the population, this prevalence is set almost entirely by the non-carrier potential outcome: P(*Y*(0) = 1) = 0.290, composed of the 20% Always-type fraction plus the remaining 80% scaled by the mean of the background risk logistic, E[expit(logit(0.10) + log(1.8)·X)] = 0.112. Carriers contribute 0.002 to the marginal prevalence. *G* has no separate effect-size parameter; its causal contribution is determined by the 40% Necessary fraction, giving P(*Y*(1) = 1) = 0.645 and a marginal PN of 0.554 (Table S3).

Figure 3A shows PN declining across the polygenic distribution. Analytic PN falls from PN(−2.25) = 0.634 to PN(2.25) = 0.397. Empirical PN falls from 0.654 to 0.439 over the same range, and GLM PN from 0.657 to 0.438. Oracle PN falls from 0.661 to 0.338; at the sparsest retained bins its binomial standard error is approximately 0.06. The 95% bootstrap interval on GLM PN contains the analytic value in the eight bins from x = −2.25 to 1.25. The variant odds ratio implied by the interaction model declines across the same range, from 5.45 at *X* = −2 SD to 3.70 at *X* = +2 SD, a reduction of 32.1%, with β_G×X_ = −0.097 (p = 0.0009). Bin-level empirical odds ratios computed from observed proportions range from 6.50 to 3.60 and are not monotone in *X* (Table S4). The 95% bootstrap interval on the empirical odds ratio contains the model-implied value in all ten bins.

**Figure 3.**
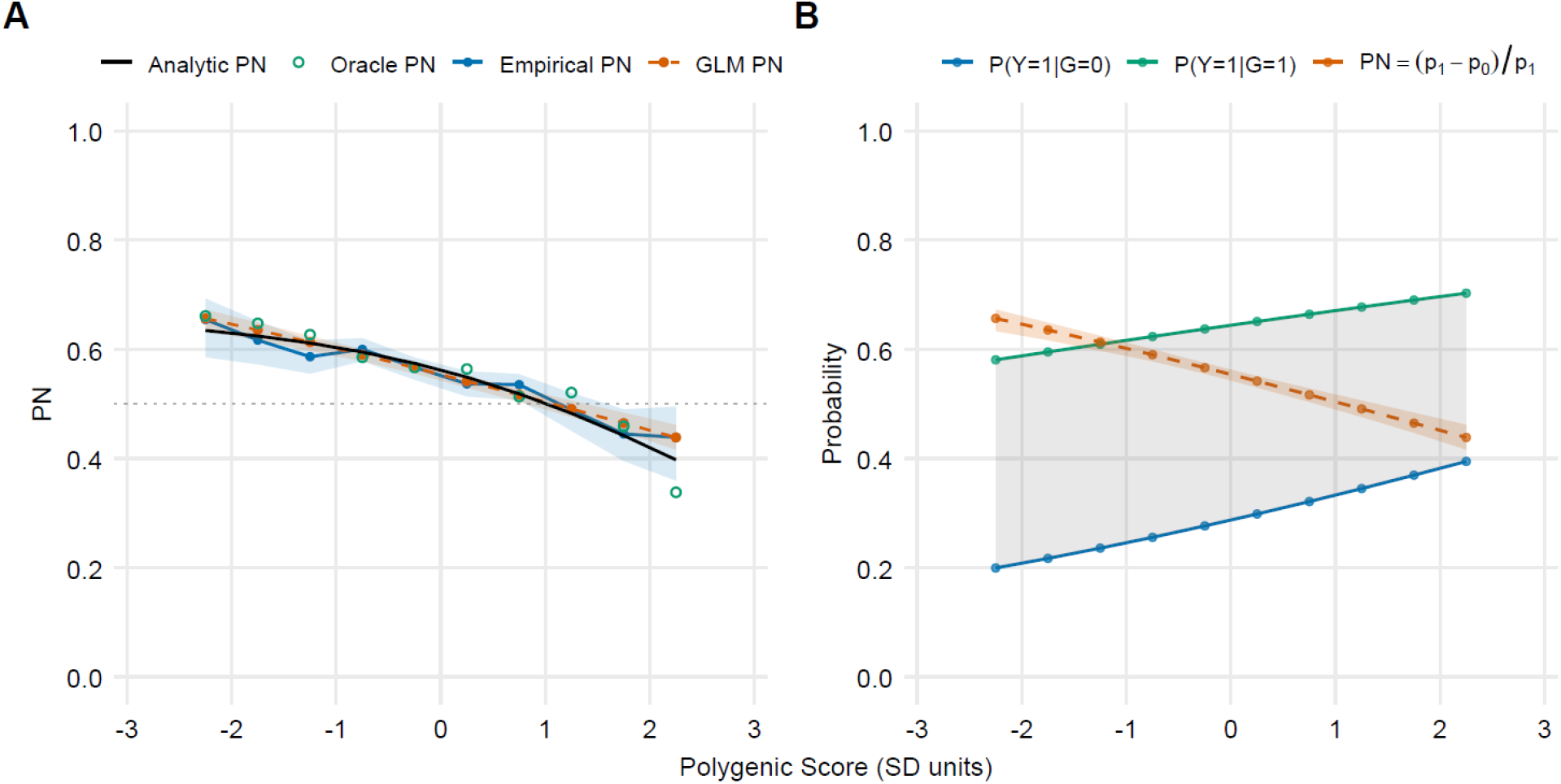
Probability of necessity declines across polygenic background in the presence of alternative causes. Simulated cohort of N = 10⁶ with independent variant carrier status G (carrier frequency 0.005) and standardized polygenic score X. Each individual was assigned a latent causal type, Necessary (0.40), Always (0.20), or Never (0.40), with monotonicity enforced. Background risk follows P(Y=1 | G=0, X) = expit(logit(0.10) + log(1.8)·X), i.e. OR = 1.8 per SD. Overall outcome prevalence was 0.291. X was divided into 20 fixed-width bins of 0.5 SD; the 10 bins containing at least 50 carriers are shown. All intervals are 200-replicate percentile bootstraps; no estimate or bootstrap replicate fell outside [0,1]. (A) Ground truth and three estimators of PN among exposed cases, PN(X) = P(Y(0)=0 | Y(1)=1, G=1, X). Analytic PN (black) is the closed-form value implied by the type mixture and the background-risk logistic, and carries no Monte Carlo error. Oracle PN (green, open circles) is the realized proportion with Y(1)=1 and Y(0)=0 among affected carriers in this simulated cohort; it is the finite-sample counterpart of the analytic curve. Empirical PN (blue) is the bin-specific plug-in (p̂₁−p̂₀)/p̂₁ from observed proportions, with a bootstrap resampling individuals within each bin. GLM PN (orange) is the same contrast evaluated at predictions from a logistic model with a G×X interaction fitted to the full sample; its bootstrap resamples the full sample and refits once per replicate. (B) Component risks underlying the Panel A decline, all from the fitted logistic model. Blue: baseline risk P(Y=1 | G=0, X). Green: carrier risk P(Y=1 | G=1, X). Grey band: the absolute risk difference between them. Orange: PN with its bootstrap interval, the same series plotted in Panel A

Figure 3B shows the fitted component risks, the penetrance and baseline. Both rise with *X*, but not equally: baseline risk P(*Y*=1 | *G*=0, *X*) rises from 0.199 to 0.395 across the retained range, an increase of 0.196, while penetrance P(*Y*=1 | *G*=1, *X*) rises from 0.581 to 0.703, an increase of 0.122. The absolute difference p₁ − p₀ narrows monotonically from 0.382 to 0.308 while p₁ rises, so the ratio (p₁ − p₀)/p₁ declines. The fitted interaction coefficient in the full model is negative, β_G×X_ = −0.097, a property which results from the non-linear relationship between the risk and log-odds scale. Table S6 reports the composition of affected carriers by assigned causal type across *X* strata. The proportion of Never-type individuals rises from 2.8% to 11.9% while the Necessary and Always proportions vary without trend around fixed expected values, since neither depends on *X*.

### Simulation Study of Biomarker-Stratified Causal Attribution

Physicians commonly measure biomarkers in addition to ordering genetic tests. The presence of biomarker data relevant to the functional impact of genetic variation can affect the causal attribution of the variant. We pursued the impact of biomarker information through the Probability of Mediated Necessity, and we performed simulations to understand its properties. At baseline simulation parameters (M-Never Always-type mass *s* = 10%, background disease rate = 5%), oracle PN was [0.58] and oracle PMN was [0.67], with the M = 1 stratum enriching for M-Caused/Y-M-only types and nearly eliminating M-Never types from the conditioning set. The PMN > PN difference reflects entirely a compositional reweighting: M = 1 conditioning collapses M-Never phenocopies (from ∼31% to ∼1% of cases), enriches high-necessity M-Caused types (from ∼54% to ∼77%), and retains M-Always types at roughly constant frequency (∼15% to ∼21%); per-type necessities are unchanged (Figure S2).

Figure 4A shows the M-Never Always-type mass sweep. As *s* increased from 5% to 60%, oracle PN eroded from [∼0.65] to [∼0.47] while oracle PMN remained stable at [∼0.72–0.74], widening the PMN − PN gap from [∼0.07] to [∼0.27]. The GLM estimator underestimated oracle PMN (conservative bias), with the gap growing with *s*. The mechanism is M-Always contamination of the G = 0, M = 1 reference cell: as *s* rises, more M-Never/Y-always phenocopies enter the M = 1 stratum among non-carriers, inflating the counterfactual risk term and causing the GLM to understate mediated necessity. The background disease burden sweep (Figure 4B) presents the contrast: as background rate increased from 0% to 30% at fixed *s*, oracle PN and PMN both decreased steeply and in near-parallel, and the oracle-GLM gap remained small and approximately flat. The divergent behavior of the two axes establishes that PMN’s robustness is specific to M-mediated Always-type mass structure, not a general insensitivity to background disease burden.

**Figure 4.**
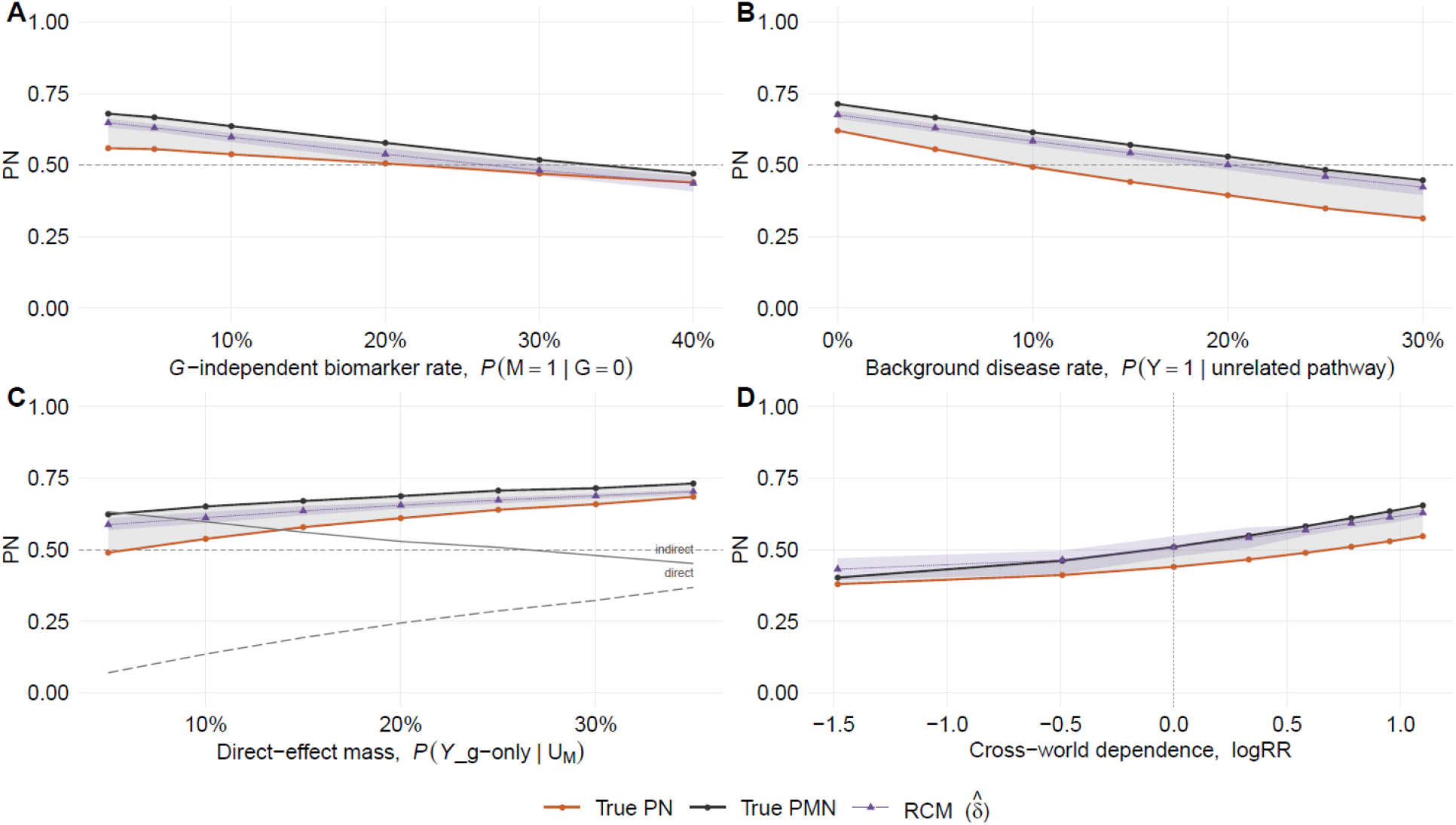
Performance of PMN Compared to PN with Incorporation of a Binary Causal Mediator. Oracle PN (orange) and PMN (grey), computed from the causal types. The RCM estimator δ̂ (purple, dashed), computed from observed with 95% interval. A. Biomarker specificity. Simulating G-independent mediator elevation, P(M = 1 | G = 0) addresses whether conditioning on M = 1 carries information about pathway activation. Increasing the proportion of individuals whose mediator is elevated by causes other than G, polygenic burden among them, so that M = 1 reflects progressively less about variant-mediated pathway activation. B. Baseline risk. Addresses the effect of alternative causes that raise the baseline risk of disease Y. The simulations raise P(Y = 1) as a uniform floor on disease probability from pathways entirely unrelated to G. C. Direct effect. Addresses the effect of increasing the non-mediated effect of G on Y, that is the direct effect. The simulations vary the mass of the Y-g-only causal type, for which disease occurs if and only if the variant is present and M is causally irrelevant. The two grey lines, labelled direct and indirect, give the causal-type composition of the PMN conditioning set on the same 0 to 1 scale as the estimands. D. Cross-world dependence. The parameter τ interpolates the joint distribution of mediator type and outcome type between independence, at τ = 0, where RCM assumption (9′) holds exactly, and the baseline coupling table at τ = 1; negative values reverse the sign of the dependence. The marginal outcome-type distribution is held fixed at every point and mediator monotonicity holds pointwise along the whole axis, so cross-world dependence is the only quantity that varies. The axis is plotted as log RR = log[P(Y-Always | M-Always) / P(Y-Always | M-Never)]. Because M(0) and M(1) are coupled along this axis, the plotted true PN and PMN are the counterfactual quantities.

It is not always clear whether a biomarker is also a causal mediator raising the risk of model misspecification and erroneous calculation of PMN. Three possible data-generating processes (DGP) involving {*G,M,Y*} were investigated: *G → M → Y* (mediated, DGP 1), *G → M* and *G → Y* with no *M → Y* effect (proxy, DGP 2), and *G → Y* and *M → Y* with *G* ⊥ *M* (independent, DGP 3). These share identical P(*Y* = 1 | *G*, *M*) cell probabilities, making them observationally indistinguishable from marginal statistics alone. However, one may examine the variable associations to try to infer whether the mediated path is more likely. A two-test protocol was used to characterize each of the three DGP. The a-path test is OR(*G, M*) in the unselected cohort, testing whether the variant is associated with the biomarker. The b-path test is defined by OR(M, Y | G = 0) i.e. testing whether the biomarker independently associates with the outcome among non-carriers; conditioning on G = 0 removes the variant’s direct effect so the residual association is attributable to M. Both pass at OR > 2. In DGP 1, both pathway tests passed [a-path OR = 4.9, b-path OR = 10.8]; the GLM PMN estimator tracked oracle PMN [∼0.71], and PMN exceeded PN [∼0.61], consistent with mediated necessity. In DGP 2, the a-path passed but the b-path failed [OR ∼1.0]; PMN was inflated by proxy enrichment; M = 1 conditioning amplifies the *G* causal signal. In DGP 3, the a-path failed [OR ∼ 1.0] and the b-path passed; PMN is arithmetically defined [∼0.45] but structurally uninterpretable as mediated necessity because *G* does not cause *M*. The two-test protocol correctly identified the structural violation in each non-mediated DGP and is a necessary, though not sufficient, condition for causal interpretability of PMN. The three DGP matched simulation (Figure S3) demonstrates when the PMN formula is and is not causally interpretable.

### Application of the PN in Ischemic Heart Disease

Genetic contributions to common diseases can include monogenic mechanisms. However, the clinical interpretation can be difficult because of incomplete or unknown penetrance, and the multiplicity of other possible causal factors. We investigated this scenario using real world data on Ischemic Heart Disease and the well-known risk produced by rare *LDLR* variants. We used data from the UK Biobank as a representative cohort not selected on disease or variant status. The analytic sample comprised 1844 heterozygous *LDLR* rare variant carriers and 488260 non-carriers after exclusions (Table 1). Among carriers, 344 met the primary IHD definition (18.7% carrier IHD rate versus 11.8% in non-carriers; OR*_LDLR_* = 1.715, 95% CI 1.525 to 1.929). There were 128,215 individuals with a recorded E78 diagnosis at any time, of whom 984 were carriers of *LDLR* rare variants. Among cases, 287/344 (83.4%) of carriers had an E78 code at any time, compared with 36,894/57,583 (64.1%) of non-carriers.

### Computational Scoring and PN Stratification within ClinVar Classes

To assess whether PN provides resolution within the VUS class, affected PN for *LDLR* variant carriers in IHD was computed stratified jointly by ClinVar classification and AlphaMissense score tertile (Figure 5A). AlphaMissense was selected as the primary scoring instrument because it derives deleteriousness predictions from protein language model representations of sequence and evolutionary context, without reference to clinical variant classifications, eliminating circularity with the ClinVar labels used as the cross-validation target. Three additional scores (ESM-1b, CADD, and REVEL) are shown alongside AlphaMissense (Figure S4) for comparison. Because PN cannot be estimated for singleton variant carriers, G was defined as carrier status within each ClinVar × score tertile stratum, and individual-level affected PN scores were computed for affected carriers within each stratum using the marginal standardization formula described above. Within-stratum spread in PN_i_ reflects variation in background IHD risk across carrier age, sex, and ancestry, not heterogeneity across individual variants.

**Figure 5.**
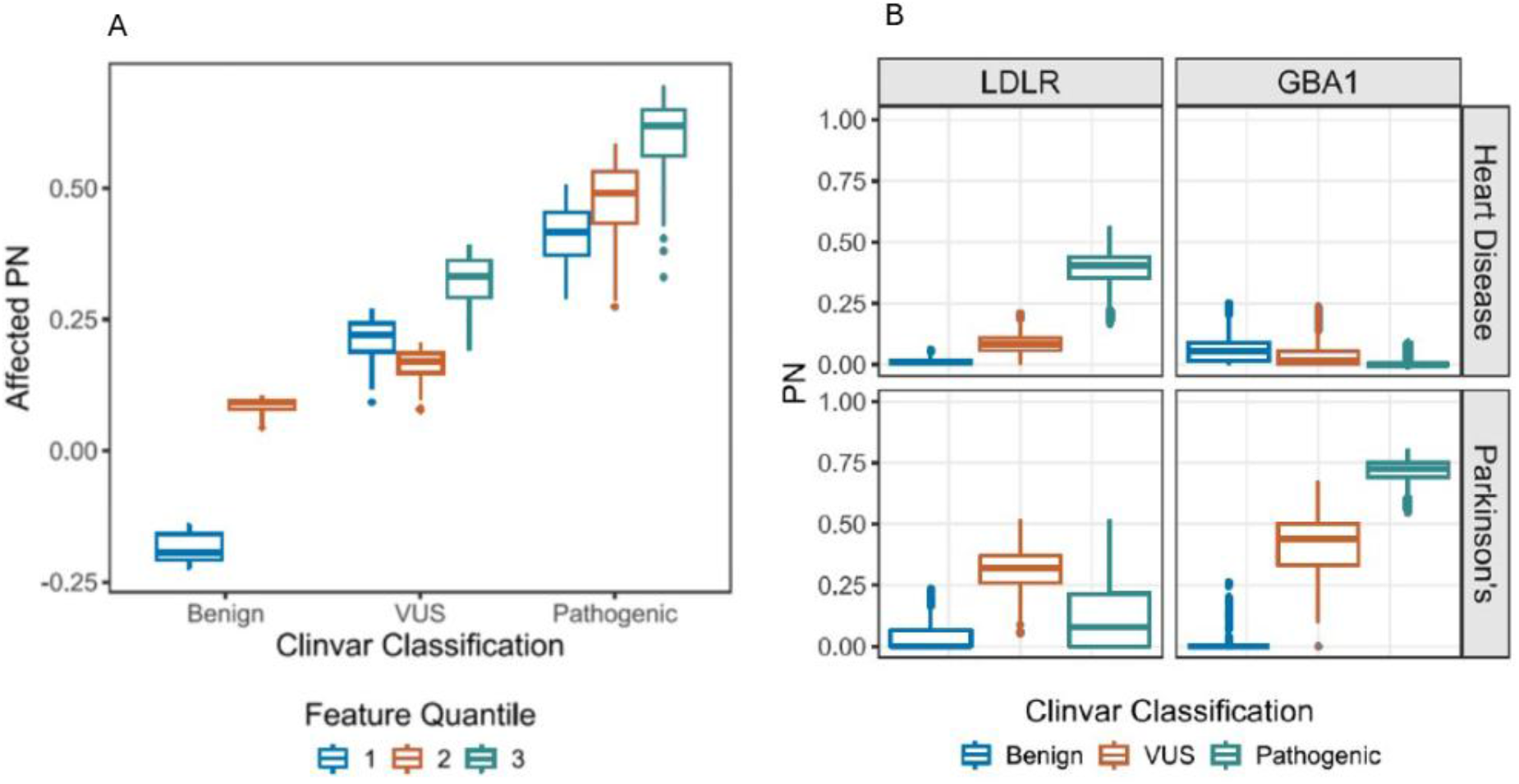
Variant Pools Defined by Deleteriousness and ClinVar Classifications. A. Composite Deleteriousness Score Effect on PN Ordered by ACMG/AMP Classification. B. Phenotype Specificity of PN

Two findings emerge from the AlphaMissense analysis. First, within the VUS class, affected PN exhibited a monotonic dose-response with score tertile: VUS tertile 1 was near zero, indistinguishable from benign variants, while VUS tertile 3 reached approximately 0.25, approaching the range of classified pathogenic variants in tertile 1. This suggests that AM score helps to resolve mechanistic heterogeneity within the VUS class. Second, within the lowest AM tertile, classified pathogenic variants had affected PN of approximately 0.30, substantially exceeding VUS variants in the same score range. Because both groups have comparable AM scores, this gap reflects information carried by ClinVar P/LP classification that AM does not capture, most plausibly functional evidence from biochemical assays, segregation, or repeated case observations that operate orthogonal to sequence-based missense scoring. AM score and ClinVar classification are therefore partially independent information sources about variant effect. PN, estimated entirely from population outcome data without using either source as an input, reflects the causal consequence from epidemiologic data, an orthogonal evidentiary signal.

The supplementary four-score comparison (Figure S4) shows that this pattern is consistent across ESM-1b, CADD, and REVEL, with AlphaMissense and ESM-1b producing the cleanest signal given their independence from ClinVar training labels. Within the Pathogenic class, ESM-1b shows less between-tertile spread than AlphaMissense, suggesting it is more informative for VUS stratification specifically than for discriminating within an already-classified population.

### Cross-Disease Validation: GBA1 Variants and Parkinson’s Disease

To assess generalizability, the ClinVar × score tertile analysis was replicated in *GBA1* variant carriers with Parkinson’s disease as the outcome. *GBA1* encodes glucocerebrosidase; heterozygous *GBA1* variants confer Parkinson’s disease risk through lysosomal dysfunction and α-synuclein aggregation^21^, a mechanism entirely independent of the LDL pathway examined in the *LDLR*/IHD analysis. Heterozygous *GBA1* carriers show approximately 5–7-fold enrichment for PD relative to non-carriers^22^, with population-based estimates placing cumulative lifetime PD risk in carriers at approximately 7–15% against a general population background of approximately 1–2%; penetrance well below the threshold at which heterozygous variants are conventionally classified as disease-causing rather than susceptibility risk factors. The Benign < VUS < Pathogenic ordering of affected PN by ClinVar class, and the monotonic tertile dose-response within each class, was replicated across all four scores in the *GBA1*/Parkinson’s system (Figure 5B). Replication across distinct biological mechanisms and organ systems supports the interpretation that the PN ordering aligns with the expected properties for estimates of causal necessity.

### Comprehensive Scoring of All *LDLR* Variant-Case Pairs

We used diagnosis of hyperlipidemia (E78 assigned prior to IHD) in UKB to evaluate PMN in the context of IHD and *LDLR* variants. Before interpreting PMN as evidence of mediated necessity, we confirmed the two required pathway tests. The a-path test (the association of variant carrier status with E78) yielded OR(*G, M*) = 3.25 (95% CI 2.96 to 3.56), confirming that *LDLR* rare variant carriers are substantially more likely to receive an E78 diagnosis than non-carriers. The b-path test (the association of E78 with IHD among non-carriers only) yielded OR(*M, Y* | *G* = 0) = 6.72 (95% CI 6.60 to 6.84), confirming that E78 is an independent predictor of IHD in the absence of rare variants. Both tests being non-null are required for a mediated interpretation of PMN.

Tier-stratified PN and PMN estimates from the training cohort are shown in Figure 6. PN as estimated here is a monotone transform of the carrier to non-carrier risk ratio, PN = 1 − p̂₀/p̂₁. Because each bin’s working dataset draws non-carriers from the same cohort at a fixed 100:1 ratio, p̂₀ is close to cohort background risk in every bin, so between-bin variation in PN is carried almost entirely by p̂₁. The tier ordering of PN restates the tier ordering of carrier risk on the risk-attribution scale. Bin-level p̂₁ and p̂₀ are given in Table S13. Where R is the ratio of the proportion of carrier cases with antecedent E78 to the same proportion among non-carrier cases, the corresponding statement for PMN is 1 − PMN = (1 − PN)/R, so PMN − PN = (1 − PN)(1 − 1/R). R is the only quantity derived from the mediator that enters PMN, and the excess of PMN over PN is bounded above by 1 − PN. Estimates followed the expected monotone gradient from higher- to lower-consequence tiers. High-confidence LoF variants (T1/F1) had the highest PN = 0.64 (∼0.50 to ∼0.71) supporting the inference that the variant was more likely than not necessary for IHD in these individuals. Strong concordant missense variants (T2/F1) yielded PN = 0.40 (95% CI 0.28–0.50), placing the point estimate below the 0.50 threshold, though the upper confidence bound reaches it. Discordant missense variants (T2_disc_/F1) had substantially lower and less precise estimates (PN = 0.21, 95% CI 0.00–0.34), reflecting both the weaker computational evidence supporting pathogenicity and the smaller carrier count in this stratum. The T3T4_low_ stratum expected value is zero under the prespecified expectation that low-evidence variants carry no causal signal for IHD, and the observed value of −0.01 (95% CI −0.06 to 0.03) is consistent.

**Figure 6.**
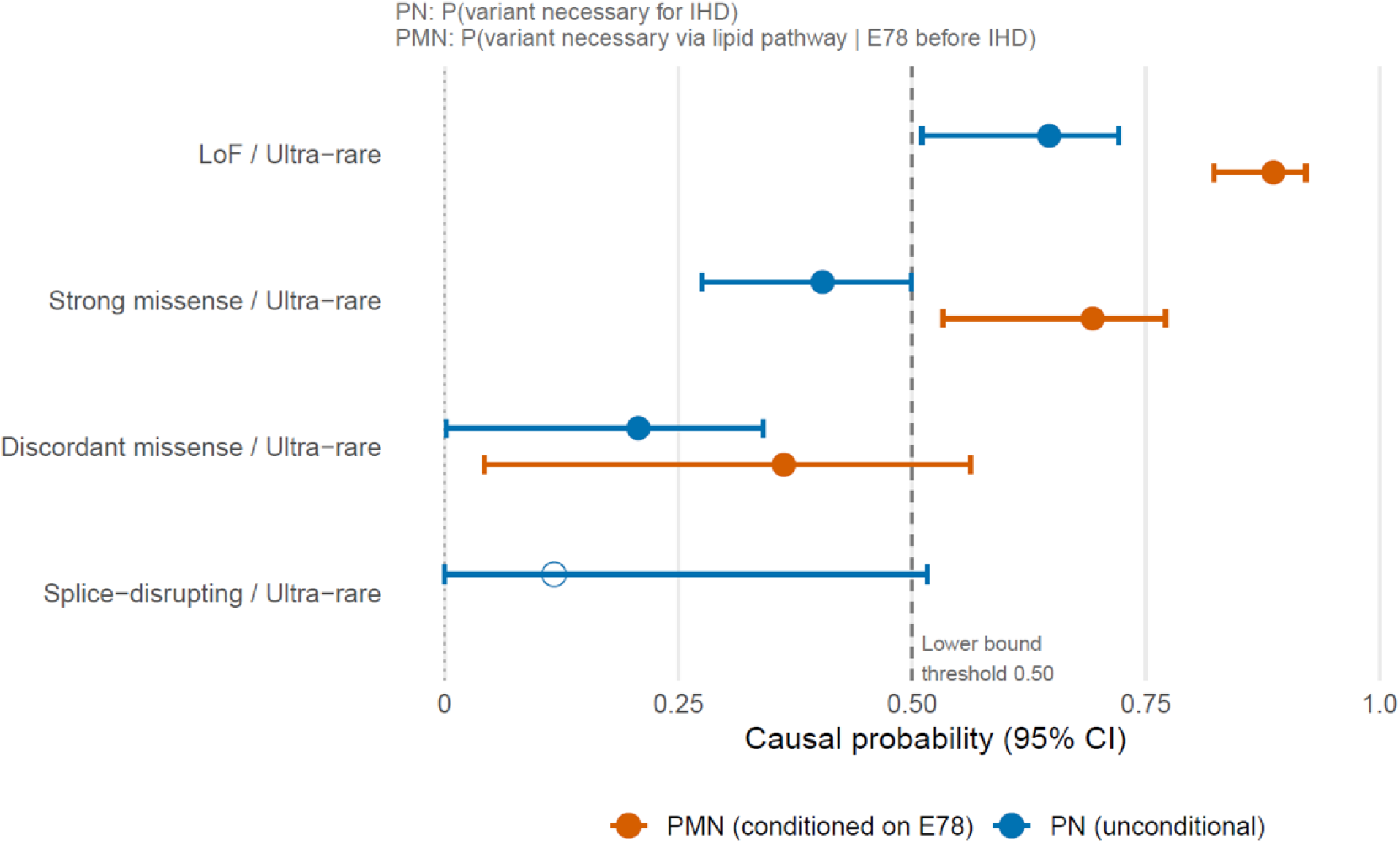
Calibration of LDLR Deleterious Rare Variants. Bin-level probability of necessity (PN, blue) and probability of mediated necessity (PMN, orange) for ischemic heart disease among heterozygous carriers of rare LDLR variants in the UK Biobank. Each row is a variant bin defined jointly by consequence tier and gnomAD v4 non-Finnish European allele frequency tier; all bins shown are ultra-rare (F1, AF < 0.01%, with variants absent from gnomAD assigned to F1). Consequence tiers were assigned from VEP sequence ontology term, SpliceAI, and computational deleteriousness scores by a prespecified script, without reference to ClinVar: LoF is T1 (loss-of-function sequence ontology term with a confirmed coding consequence); strong missense is T2 (at least two strong votes among REVEL ≥ 0.75, AlphaMissense ≥ 0.80, ESM-1b ≤ −7.5); discordant missense is T2_disc (exactly one strong vote); splice-disrupting is T_splice_ (SpliceAI maximum delta ≥ 0.50, applied as an override to any sequence ontology term). Carriers of variants in more than one bin were assigned to a single analysis bin by a prespecified consequence priority order. For each bin a working dataset was formed from all carriers in that bin together with one random subsample of non-carriers at a 100:1 ratio, drawn once prior to analysis. Two logistic models with identical linear predictors were fitted to that dataset: a primary outcome model for IHD and a joint outcome model for the composite event (IHD with E78 recorded before IHD). Both included age at recruitment, sex, a standardized CAD polygenic risk score, and the first four ancestry principal components. PN and PMN were formed by marginal standardization, setting G = 1 and G = 0 for every individual while retaining observed covariates, averaging the predicted risks across the working dataset, and taking one minus the ratio of the G = 0 to the G = 1 average. Estimates are therefore marginal over polygenic background rather than conditional on it. Intervals are 95% bootstrap percentile intervals from B = 1000 resamples of the pre-thinned working dataset, refitting both models on each resample. Filled points denote bins meeting the prespecified estimability criteria for both models; the open point denotes an estimate affected by complete separation in the non-carrier stratum, for which the point estimate is shown without an accompanying PMN. The vertical dashed line marks the prespecified 0.50 reference value; the dotted line marks zero.

PMN exceeded PN in all tiers with sufficient E78 prevalence to support estimation. The PMN = 0.89 (∼0.79 to ∼0.91) for T1 /F1 and 0.69 (∼0.59 to ∼0.81) for T2/F1, reflecting the enrichment for Necessary-type individuals when analysis is restricted to carriers with antecedent hypercholesterolemia. This is consistent with the simulation finding that PMN exceeds PN when the G → M → Y pathway dominates and M is specific to the genetic mechanism.

Of the 22 withheld IHD cases carrying an *LDLR* rare variant. The majority of holdout carriers in the T2/F1 and T1 /F1 strata had antecedent E78. PRS tertile distribution was spread across low, mid, and high strata within each bin, with no systematic enrichment for high PRS in higher-consequence bins (data not shown).

## Discussion

When a clinician names a cause for a patient’s disease, whether a drug, an environmental exposure, an infection, or a genetic variant, the claim is counterfactual: that without the named antecedent, this patient would not have this disease. We have explored a framework for causal assessment of germline variation under which such a claim can be evaluated quantitatively, albeit probabilistically. Unlike an exposure that must be recalled or bacteria cultured after the fact, the candidate causal variant is fixed at conception, measured directly, and observable at population scale in cohorts assembled without regard to the disease. What makes the calculation challenging is not the number of variants a patient carries but how often the phenotype occurs in their absence. That burden is heaviest for phenotypically broad conditions with complex causal architectures, which include late-onset common diseases but also intellectual disability, epilepsy, congenital anomalies, and autism, where testing has long been standard and attribution has been reported as diagnostic yield. We have treated the case as the unit of analysis treating the variant(s) as the exposure, and we developed molecular diagnosis as a probability of necessity (PN). Our results argue that molecular PN is identifiable from population data under clearly articulated assumptions. From this vantage, molecular diagnosis is a causal statement dependent on competing risks and mediating pathways present in the population from which it is estimated, and it should be adjusted according to the specific context and attributes of the patient. Under the PN framework, genetic test interpretation and variant attribution are not a fixed properties of a genetic variant(s), but instead should be viewed as a probability that aligns to the circumstances, source population and context of the patient in which genetic testing has been applied.

We focus on PN rather than probability of sufficiency (PS) or probability of necessity-and-sufficiency (PNS) because the minimum requirement of diagnosis is identification of an active causal pathway such that the patient would not have had the disease but for the presence of the variant. PN estimates operate at the diagnostic thinking level of clinical utility^19^, estimating certainty that a molecular diagnosis has been achieved. Current clinical utility studies of genetic testing implicitly assume molecular diagnoses are accurate when a variant classified as pathogenic is identified, but this approach depends on an informal exclusion principle where the realized probability varies across phenotypes that differ in prevalence and alternative causes. By scaling diagnostic certainty as a probability, PN could provide a stronger foundation for assessing both case interpretation for individuals as well as clinical utility of the test procedure itself.

### PN-Penetrance Dissociation

The dissociation of penetrance and PN is a key finding of our work. Penetrance is a foundational concept in human genetics, but it is known to be complicated by confounding and selection bias. PN enables baseline disease risk and competing causes to enter into diagnosis-based decision making. The *GBA1*/PD results from UKB demonstrate this idea empirically; the high PN values of *GBA1* variants follow directly from the structure of the estimand. PN = (p₁ − p₀)/p₁. The ratio of excess carrier risk to total carrier risk among cases is algebraically identical to the etiologic fraction among the exposed, the classical epidemiological quantity measuring the proportion of disease burden in exposed individuals that is attributable to the exposure.^9^ PN is governed by the relative contrast between carrier and non-carrier risk, not by variant penetrance alone. A variant that elevates disease risk several-fold from a low baseline prevalence produces high PN among affected carriers even when penetrance is low, because the carrier-attributable excess is large relative to background. This is the configuration of *GBA1* in PD. A variant can fail to have high penetrance while being strongly implicated as the but-for cause of disease in affected carriers even when accounting for covariates, and this can meaningfully inform management and clinical thinking.

### Effect of Patient Selection

Estimation of PN requires frequency information on the co-occurrence of causative alleles and disease phenotypes. These data must be obtained by observation, but not everyone in a target population typically receives genetic testing. The prioritization of individuals for testing can strongly impact PN estimates. In our simulation studies, selection acts on disease status and, conditional on disease status, is independent of causal type., The true underlying causal-type composition of affected carriers is therefore indistinguishable from that under an unselected testing regimeHowever, in a heavily ascertained cohort where non-diseased individuals are excluded bias in naïve estimation of PN arises from the comparison group. Because patients without disease are referred for testing far less often than patients with it, selection multiplies the odds of disease among both carriers and non-carriers by the same factor, λ. The lower non-carrier risk rises proportionally more than the carrier risk, compressing the excess-risk ratio from which PN is computed and biasing PN downward. Disciplined clinical practice narrows referral itself to patients with the phenotype under study, so a comparably referred, phenotype-negative population rarely exists in clinic-ascertained cohorts to serve as a denominator for baseline risk. Baseline risk is typically absent from the reported cohort in most clinic-based diagnostic yield studies. This phenomena is directly analogous to the depression in polygenic risk association to disease that is observed in a clinically selected population compared to the general population.^23^

Correction for selection bias is possible in principle. The bias depends only on how much more frequently a patient with disease is referred for testing than a patient without, and when that ratio is supplied and used for adjustment, our simulations recover the true PN exactly. The ratio can be estimated by comparing the tested cohort with the population from which it was drawn. The correction assumes that referral depends on disease status alone. Where referral also depends on family history or prior genetic testing, selection depends on genotype directly, and selection through unmeasured severity poses the same problem; neither is addressed by this correction. This is an area for future research.

Given the issues with clinic-based samples, we therefore recommend that PN and related measures be initially estimated from population-based biobanks (e.g., UK Biobank, All of Us), where recruitment is driven by demographic rather than disease factors and the ratio of cases and non-cases is preserved, albeit with age, healthy-volunteer, and other potential selection biases.

### PN in the Presence of Alternative Causes

Our simulations show that independent alternative causes produce systematic heterogeneity in PN. When a strong alternative pathway to disease is present, a given variant is less likely to have been necessary for an individual case: at low alternative-cause burden most affected carriers needed their variant, whereas at high burden many would have become affected regardless. Because PN is defined among cases, it conditions on disease status, a common effect of the variant and the alternative cause; this is the collider structure we exploited in formulating the Causal Pivot.^24^ The gradient therefore follows from the structure of the estimand and does not require biological interaction between the variant and the alternative cause. The same structure separates PN from penetrance. More carriers in high-burden strata would have developed disease without the variant, so penetrance rises with alternative-cause burden while PN falls. This bears directly on interpreting PN in multifactorial disease, where polygenic scores provide a measured alternative cause.

### Mediated Necessity

In this study we examined the canonical scenario where there is additional information about a causal mediator represented by *G→M→Y*. Physicians regularly use such information to confirm that a putative variant-driven causal pathway is active in an individual, and the pursuit of such biomarkers is crucial to molecular medicine. To formalize the contribution of biomarker data to diagnostic certainty in genetic diagnostics, we the statistic PMN which conditions on the mediator value, and we extended our simulation framework to account for the new levels of causal types. We found the conditions under which PMN > PN are a structural consequence of the conditioning set (*M*=1). M-Never individuals (*M*(1) = 0 makes *M*=1 unachievable for this causal type) cannot contribute to the *G*=1, *M*=1, *Y*=1 stratum. Among M-Never individuals, those who develop Y through pathways entirely independent of *M* (the M-Never × Y-Always subclass) appear in the plain PN conditioning set as phenocopies contributing nothing to individual necessity, because their disease has no dependence on either *G* or *M*. The simulation confirms that the PMN exceeds PN because the target population has changed. The residual phenocopies (M-Always × Y-Always) remain in the PMN set and bound how far PMN can exceed PN. We also demonstrated ordered testing for associations between the three variables establishes the necessary preconditions before PMN is interpreted as mediated necessity. From a clinical perspective, these findings support the use of mechanistic biomarkers to refine variant pathogenicity assessment. A patient presenting with both a candidate variant and an elevated pathway-specific biomarker is more likely to be explained by *G* than an equivalently affected patient without biomarker elevation. This stratification informs both diagnostic interpretation and therapeutic decision-making where targeted interventions impact the mediated pathway.

### Ischemic Heart Disease

Because causal variants are typically ultra-rare or private, no per-variant estimate is readily available and the exposure *G* must be defined on a variant pool or class. How that pool is constructed determines the interpretation of what PN measures. PN is estimated within the stratum of the population the pool defines, so if membership depends on which carriers were affected, the carrier risk r₁ is inflated and the estimate is circularly dependent on the evidence that built the variant class. Variant class membership should therefore not be a function of the disease outcomes in the study sample used for estimation. This constrains the available annotations that should be used in establishing variant classes. Allele frequency and sequence-based deleteriousness scores are computed from population and protein data without consulting the phenotypes of the individuals under study* (except survival), whereas clinical labels and association statistics are functions of observed outcomes. We constructed pools using allele frequency and loss of function annotation and AlphaMissense, and the latter was replicated with ESM-1b, an independently trained variant score. The cost is that pools are variably contaminated with neutral variants, which attenuates r₁ toward r₀ and lowers pool-level PN relative to the value for a truly deleterious member. That attenuation is conservative, and we prefer it to an estimate whose value may in part be circular with the evidence used to select the variants.

We applied our methods to real data from UKB. Our objective was both to confirm the methodology as well as to examine its potential to resolve otherwise ambiguous case-variant pairs. We chose ischemic heart disease as a demonstration phenotype focusing on the diagnostic utility of variation in *LDLR*. Ischemic heart disease is a complex phenotype with multiple etiologic components including lipids and other pathways.^25,26^ The dose-response between AM score tertile and PN within the VUS class, and the persistent PN advantage of classified pathogenic variants over VUS at equivalent AM scores, are jointly explained by the partial orthogonality of the three evidence dimensions the analysis brings together. AlphaMissense operates on protein sequence and structural context; information that is variant-intrinsic and computable without clinical observation. ClinVar classification synthesizes heterogeneous evidence: previous observation in an affected person, functional assay data, co-segregation in pedigrees, case-series enrichment, and loss-of-function annotations. These are sources that document biological consequence through routes that sequence-based scoring does not access and that accumulate over years of clinical observation. PN is estimated from population outcome frequencies without direct reference to either source, capturing the causal consequence of whichever molecular mechanism is operating. The dose-response within VUS reflects AM’s capacity to detect functional consequence. Our results suggest that PN, derived from epidemiologic evidence (excess relative risk) independent of variant-level annotation, orders VUS-classified cases by predicted causal necessity without altering ACMG/AMP classification of the variant. A case ordered high by PN remains a VUS; PN adds case-level information that variant classification alone does not provide.

### Parkinson’s Disease and *GBA1*

We also examined PN in the context of PD and variation in *GBA1.*^22,27^ On the one hand, *GBA1* carrier status is known to have incomplete penetrance. On the other hand, approximately 10-15% of European PD cases are *GBA1* carriers. We sought to ask the question of diagnostic attribution of *GBA1* in carrier cases. Our results indicate that *GBA1* is strongly necessary for PD among affected carriers. Although this result may seem surprising given the more modest penetrance, the PN result is strongly driven by the lower prevalence of PD.

## Limitations

Estimating counterfactual probabilities requires correctly specified models and strong assumptions. Remaining bias in the estimates of PN may be driven by violations of the underlying assumptions and the variance of estimates is driven by limitations in sample size and phenotyping. An important concern is that lifetime exposures including factors that may be considered environmental occur postzygotically, and are therefore technically post-treatment. When such exposures are downstream of the variant’s causal mechanism, conditioning or controlling for such exposures can bias PN estimates. This is an area for future research. Moreover, under some circumstances PN may not be estimable if there is insufficient overlap between available reference cohorts and the clinically relevant test population. There may also be unmeasured confounding. If there are unmeasured *U* affecting both *G* and *Y*, the prediction of baseline *G* effects will be biased. There is somewhat less concern for this in genetics after ancestry has been taken into account.

Model misspecification is a potential problem, including wrong functional form, missing interactions, and non-linearities. A non-zero G × X coefficient in a fitted logistic model does not by itself indicate biological effect modification; both p₀ and p₁ rise with X while their proportional gap narrows, producing risk saturation without a change in G’s causal effect (Results)

Finally, the quantities reported here are defined at the level of the variant pool. Covariates enter both fitted models, but the predicted risks are averaged over the covariate distribution of the working dataset before the ratio is formed, so the bin-level PN and PMN of Figure 6 are marginal over age, sex, ancestry and polygenic background. Estimates conditional on polygenic burden would require stratification on PRS, which the present cell counts do not support.

## Future Directions

Transportability of PN across populations requires decomposing differences into contributions from baseline risk and from carrier risk itself, since cohorts differing in ancestry vary in both competing-cause burden and variant-pool composition, and both bear on how variant interpretation is performed in practice. Recruitment era and design shift baseline risk independently of the variant pool, so PN estimates require periodic re-derivation rather than a single fixed value per variant. Population cohorts can also benchmark deleteriousness predictors against PN directly, avoiding the circularity of curated pathogenicity labels; we pursue this elsewhere.

Covariates occurring after the genetic exposure, which is fixed from conception, raise separate concerns. PRS, sex, and ancestry are legitimately pre-treatment; age, environmental exposures, and lifestyle variables in UKB are not. Age is particularly concerning because biobank enrollment conditions on survival to recruitment. This limits PN estimation for phenotypes such as intellectual disability and autism, which are routinely tested genomically but for which no population resource currently permits PN estimation.

Variant pools defined by continuous deleteriousness score order variants along one severity axis; pools defined by protein domain or mechanism instead partition by pathogenic process. In *LDLR*, alleles affecting ligand binding, intracellular transport, receptor recycling, and internalization differ in residual receptor function by mechanism rather than predicted severity, and mechanism-based pools should separate more sharply in PMN than in PN. A domain where necessity fails to track severity would be a direct test of monotonicity.

Formal sensitivity analysis for the cross-world independence assumption remains the principal open theoretical problem for PMN. Existing sensitivity frameworks are built for natural direct and indirect effects and do not transfer directly to necessity, where the direction of bias is not constant across the dependence axis. PN itself does not require this assumption.

## Conclusion

The probability of necessity is an established causal quantity, applied here to molecular diagnosis in the context of a variant-case pair. We argue this is both the unit of diagnostic thinking as well as the target of clinical utility. Through this formalism the causal claim in a diagnosis acquires an explicit probabilistic form, an estimator by which it can be calculated and through which its uncertainty can be assessed, and a set of conditions under which the causal interpretation holds. The quantity is estimable in existing population cohorts.

## Declarations

### Data availability

Individual-level UK Biobank data cannot be redistributed by the authors. They are available to bona fide researchers by application to UK Biobank (https://www.ukbiobank.ac.uk/enable-your-research/apply-for-access). Derived quantities sufficient to reproduce every figure and table, including bin-level carrier and non-carrier counts, event counts, fitted coefficient vectors, and variance-covariance matrices, are provided in the Supplemental Material as aggregate statistics permitted for export under the Material Transfer Agreement. Publicly available resources used are gnomAD v4 (https://gnomad.broadinstitute.org), and ClinVar (https://www.ncbi.nlm.nih.gov/clinvar).

### Code availability

Simulation code and the documents that generate Figures 1 through 4 and Supplemental Figures S1 through S3 are available at https://github.com/geneticgenomicservices/ProbNecessity. The repository contains no individual-level data and no code that operates on UK Biobank data. Analysis scripts executed within the UK Biobank Research Analysis Platform are available from the corresponding author on request, subject to UK Biobank access terms. The repository is publicly readable to support review and verification; reuse beyond verification requires permission from Baylor College of Medicine.

### Author contributions

Conceptualization, [J.W.B. and C.A.S.]; Methodology, [J.W.B., C.A.S.]; Software, [J.W.B., C.J.W.]; Formal Analysis, [J.W.B. and C.A.S.]; Investigation, [J.W.B]; Data Curation, [J.W.B., C.J.W.]; Writing - Original Draft, [J.W.B.]; Writing - Review and Editing, [J.W.B., C.J.W., C.A.S.]; Visualization, [J.W.B.]; Supervision, [C.A.S.]. All authors read and approved the final manuscript.

### Use of AI

Portions of this manuscript were drafted and revised with the assistance of Claude (Anthropic PBC), a large language model AI assistant. AI assistance was used for structural and logical review of scientific arguments, R and python code drafting, manuscript text drafting, cross-checking of methods text against analysis code, and creating a code repository. All scientific claims, analyses, statistical models, and final text were edited, verified, and approved by the authors. The AI played no role in the original conception of the PN/PMN application in genetic diagnostics. nor did it initiate or perform any original statistical analyses. All simulations and data analysis were conducted by the authors in R and, where noted, the UK Biobank Research Analysis Platform. The authors take full responsibility for the integrity and accuracy of all published content.

## Acknowledgments

This research was conducted using the UK Biobank Resource under Application ID 98786. We thank the UK Biobank participants and the staff who established and maintain the resource.

## Footnotes

† Setting *w*_1_ = *γ*_0_ in place of *γ*_0_/*γ*_1_ amounts to asserting *P*(*M*(0) = 1 ∣ *M*(1) = 1) = *P*(*M*(0) = 1), that is, *M*(0) ⊥ *M*(1). This is cross-world independence of the potential mediators.

‡ RCM additionally derive an influence-function-based estimator for a user-specified projection of *δ*(*x*) that attains the nonparametric efficiency bound.

§ Uniformity is definitional: a type whose outcome does not depend on *m* cannot be concentrated in one mediator-type stratum without making the direct route conditional on mediator behavior, which is the opposite of what direct effect denotes.

