## Supplementary Material for "Molecular Diagnosis as a Probability of Necessity"

#### Identifying Assumptions

**Table S1. Identification assumptions for PN and PMN, with correspondence to the assumption set of Rubinstein, Cuellar and Malinsky (2025).**[1]

| Assumption | RCM | Formal statement | Consequence of violation |
|---|---|---|---|
| A1 | (6) | $M = M(g)$; $Y = Y(g,m)$; no interference | Estimand undefined rather than biased |
| A2 | part of (8′) | $Y(0) \perp G \mid Z$ | Sign of bias set by sign of residual G–Y confounding |
| A3 | — | $Y(1) \geq Y(0)$ | Pearl's formula returns an upper bound; reported PN is anti-conservative |
| A4 | part of (8′) | $M(g) \perp G \mid Z$ | Biases $\gamma_0$ and $\gamma_1$, hence $w_1$; sign indeterminate |
| A5 | implied by (8′), (9′) | $Y(0,m) \perp M \mid G, Z$ | Inflates $\mu_{01}$ and $\mu_{11}$ jointly, so the two effects partly offset; sign indeterminate |
| A6 | — | No unmeasured L with $G \rightarrow L$, $L \rightarrow M$, $L \rightarrow Y$ off the $M \rightarrow Y$ path | Invalidates A4, A5 and A7 jointly |
| A7 | (9′) | $\{M(0), M(1)\} \perp Y(0,m) \mid Z$, for $m = 0, 1$ | No observable quantity changes; $\delta$ is not point-identified; $\hat{\lambda}$ is uninformative about this violation |
| A8a | (7′), mediator part | $M(1) \geq M(0)$ | $P(M(0)=1 \mid M(1)=1) \neq \gamma_0/\gamma_1$; $\hat{\delta}$ weights the wrong stratum while every observable is unchanged; sign indeterminate |
| A8b | (7′), outcome part | $Y(1,1) \geq Y(0,m)$, for $m = 0, 1$ | $\hat{\delta}$ returns an upper bound on mediated necessity |
| A9 | (10′), (11) | $P(G=g, M=m \mid Z=z) \geq \varepsilon > 0$ for all g, m, z; $P(Y=1 \mid G=1, M=1, Z=z) \geq \varepsilon > 0$ | $\hat{\mu}_{00}$, $\hat{\mu}_{01}$, $\hat{\mu}_{11}$ or the denominator of $\delta$ not estimable; $\hat{\delta}$ unstable rather than biased |

#### Causal Types and the Potential Outcomes Framework

Our base simulation framework draws on the causal types representation of the potential outcomes model (Brumback, 2021).[2] Under the monotonicity assumption $Y(1) \geq Y(0)$ , which excludes individuals for whom the variant would be protective, each individual belongs to exactly one of three latent types defined by their joint potential outcomes $\{Y(0), Y(1)\}$ (Table S2). The fourth logical combination $\{Y(0)=1, Y(1)=0\}$ is ruled out by the monotonicity assumption. These types are unobservable in real data since each individual is observed under only one exposure level. In these simulations ground-truth causal type membership is produced by construction. The 'truth' PN is then the population fraction of Necessary types among affected carriers, i.e. those with G=1, Y=1.

**Table S2 Causal Types Invoked in Simulations**

| Type | Y(0) | Y(1) | Interpretation | Contributes to PN? |
|---|---|---|---|---|
| Necessary | 0 | 1 | Disease requires G | Yes |
| Always | 1 | 1 | Disease regardless of G (phenocopy) | No |
| Never | 0 | 0 | No disease regardless of G | No |
| ~~Preventive~~ | ~~1~~ | ~~0~~ | ~~G prevents disease~~ | ~~Excluded by monotonicity~~ |

### Effect of Case Selection

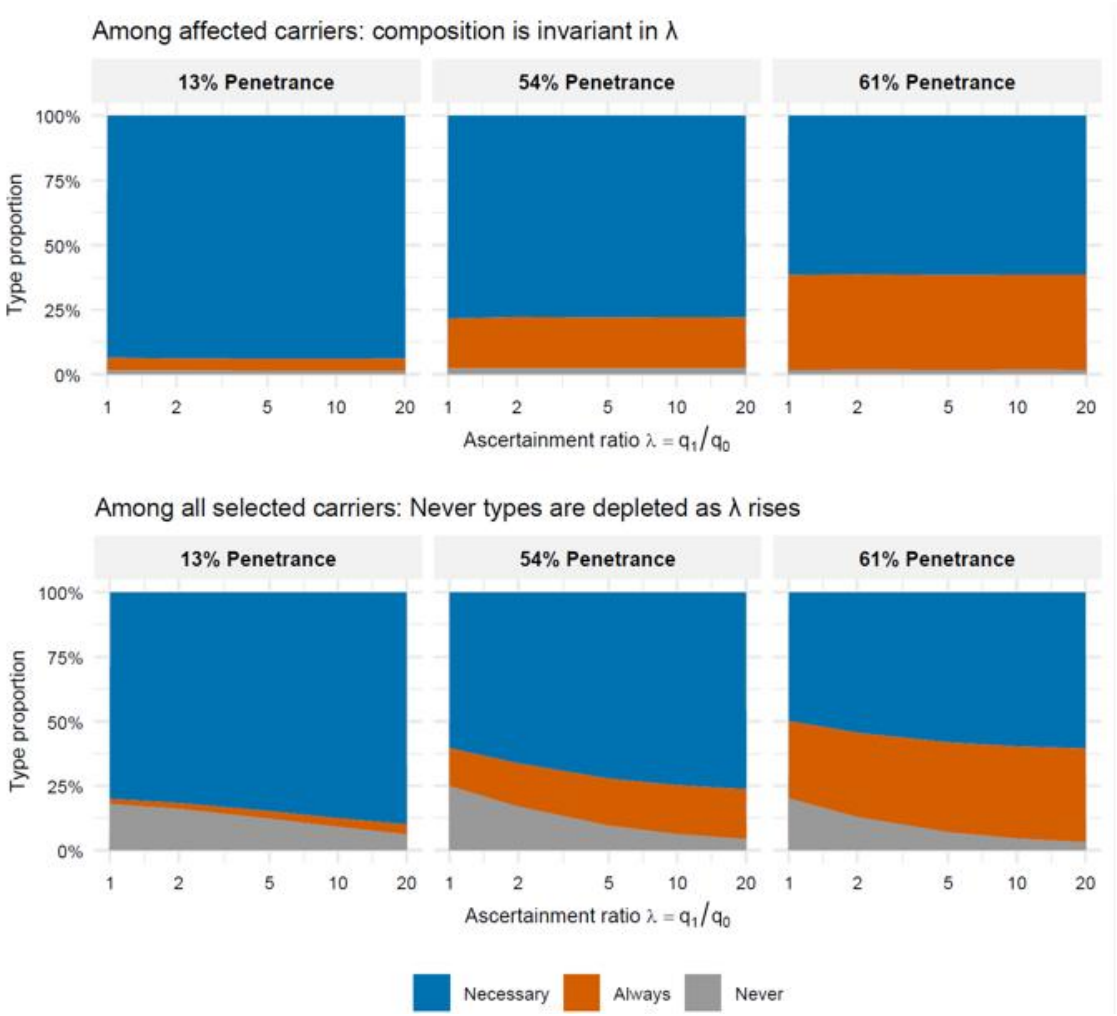


Figure S1. Causal type composition in ascertained samples, by denominator. Stacked areas show the proportions of Necessary (blue), Always (orange), and Never (grey) types as a function of the ascertainment ratio $\lambda = q_1/q_0$. Columns correspond to the three disease scenarios, labelled by penetrance; population type proportions are 80/2/18% (13% penetrance), 60/15/25% (54%), and 50/30/20% (61%) for Necessary/Always/Never. (A) Among affected carriers (G=1, Y=1, S=1), composition is invariant in λ. (B) Among all selected carriers (G=1, S=1), regardless of disease status, Never types are progressively depleted as λ rises, since they are rarely affected and therefore rarely referred. Counts pooled across 100 Monte Carlo replications ($N = 10^6$ per replicate).

Among affected carriers the type composition is invariant in λ (Figure S1A). Ascertainment acts on Y alone, so $S \perp \text{type} \mid Y = 1$ and P(type | G=1, Y=1, S=1) = P(type | G=1, Y=1) for every λ. The affected carriers in a heavily ascertained cohort are, in causal-type composition, indistinguishable from those in an unselected one. Composition differs across scenarios because types are reweighted by type-specific penetrance: Always types comprise 4.7%, 19.5%, and 36.9% of cases against population proportions of 2%, 15%, and 30%.

Among all selected carriers, composition does move with λ, but by depletion of Never types (Figure S1B). At λ = 1 it recovers the population proportions. Necessary and Always types are both enriched, Necessary more so in absolute terms, because the two share the same p_Y1 in Scenarios B and C; ascertainment on disease status cannot separate them.

Together these establish the mechanism as risk compression rather than case recomposition. Ascertainment drives both P(Y=1 | G=1, S=1) and P(Y=1 | G=0, S=1) toward 1, and the excess-risk ratio $(r_1 - r_0)/r_1$ shrinks. The bias cannot be detected from within the ascertained cohort because the individuals who were not referred are missing from the denominator.

### Alternative Causes

**Table S3**. Population summary by latent causal type. Risks are P(Y=1|G), potential outcomes are marginal means. Oracle PN is computed among G=1, Y=1 cases within each type.

| type | pop_share | p_Y_g0 | p_Y_g1 | p_Y0 | p_Y1 | pn_oracle | affected carriers |
|---|---|---|---|---|---|---|---|
| Always | 0.20 | 1.000 | 1.000 | 1.000 | 1.000 | 0.000 | 986 |
| Necessary | 0.40 | 0.112 | 1.000 | 0.112 | 1.000 | 0.883 | 2039 |
| Never | 0.40 | 0.111 | 0.110 | 0.111 | 0.111 | 0.000 | 222 |
| Total | 1.00 | 0.289 | 0.643 | 0.289 | 0.644 | 0.554 | 3247 |

Observed risks P(Y=1|G) are shown alongside marginal potential-outcome means P(Y(g)=1); their agreement confirms that G was assigned independently of causal type and of X. Oracle PN is computed among affected carriers within each type. Necessary-type carriers have oracle PN below 1 because a Necessary-type individual for whom the alternative cause is also active has Y(0) = 1 and contributes no necessity. The total row gives the population-average PN of 0.554, against which the polygenic-stratum values in Figure 3A range from 0.634 to 0.397.

**Table S4. Variant odds ratio by polygenic bin X**

| X | n_g1 | or_emp | or_model |
|---|---|---|---|
| -2.25 | 90 | 6.50 (4.47, 11.1) | 5.59 (5.01, 6.33) |
| -1.75 | 228 | 5.12 (3.98, 6.66) | 5.32 (4.86, 5.88) |
| -1.25 | 475 | 4.38 (3.80, 5.28) | 5.07 (4.74, 5.48) |
| -0.75 | 769 | 5.16 (4.50, 5.97) | 4.83 (4.57, 5.14) |
| -0.25 | 933 | 4.43 (3.93, 5.05) | 4.60 (4.37, 4.83) |
| 0.25 | 937 | 4.10 (3.63, 4.70) | 4.38 (4.16, 4.61) |
| 0.75 | 770 | 4.60 (3.98, 5.41) | 4.18 (3.91, 4.44) |
| 1.25 | 460 | 4.00 (3.38, 4.80) | 3.98 (3.67, 4.32) |
| 1.75 | 230 | 3.60 (2.76, 4.81) | 3.79 (3.37, 4.21) |
| 2.25 | 91 | 4.56 (3.01, 8.01) | 3.61 (3.14, 4.11) |

Empirical values are computed from observed proportions within each bin, with percentile intervals from 200 within-bin bootstrap replicates. Model values are $\exp(\beta_G + \beta_{GxX} * X)$ from the full-sample interaction fit, with intervals from 200 full-sample bootstrap refits. The two interval types are not comparable: the empirical interval reflects bin-level sampling variability, the model interval reflects coefficient uncertainty only. The 95% bootstrap interval on the empirical odds ratio contains the model-implied value in all ten bins. These intervals are wide where carriers are sparse, spanning 4.47 to 11.1 at x = −2.25 and 3.01 to 8.01 at x = 2.25, and they overlap across the full range.

**Table S5** GLM Coefficients: Full Interaction Model in Alternative Cause Simulation

| | Interpretation | Estimate | SE | z_value | p_value |
|---|---|---|---|---|---|
| (Intercept) | Intercept | -0.9091 | 0.0022 | -407.4574 | 0e+00 |
| G | G main effect (log-OR per copy) | 1.5021 | 0.0295 | 50.8365 | 0e+00 |
| X | X main effect (log-OR per SD) | 0.2183 | 0.0022 | 97.3675 | 0e+00 |
| G:X | G×X interaction (log-OR per copy × SD) | -0.0969 | 0.0292 | -3.3212 | 9e-04 |

**Table S6. Composition of affected carriers by assigned causal type across polygenic strata.**

| X Stratum | n_stratum | Always | Necessary | Never |
|---|---|---|---|---|
| (-Inf,-0.67] | 809 | 31.52 | 65.64 | 2.84 |
| (-0.67,0] | 759 | 31.88 | 62.45 | 5.67 |
| (0,0.67] | 767 | 29.47 | 62.45 | 8.08 |
| (0.67,1.35] | 600 | 26.33 | 64.17 | 9.50 |
| (1.35, Inf] | 312 | 33.65 | 54.49 | 11.86 |

Percentages are among carriers with Y = 1 in each stratum. Expected values follow from the data-generating process: the Necessary and Always proportions are 0.40/D and 0.20/D with D = 0.60 + 0.40·E[p(X)], neither of which depends on X, so their ratio is fixed at 2:1 and their variation across strata is sampling noise. The Never proportion is 0.40·E[p(X)]/D and rises with X as the alternative cause activates more often.

**Note on Figure 3B**

Penetrance, P(Y = 1 | G = 1, X), is a conditional probability rather than a causal effect. A geneticist who observes penetrance rising with polygenic score may conclude that the variant is more penetrant on a high-risk background. However, in our simulation the joint distribution of potential outcomes is held fixed across strata of X, and penetrance rises only because baseline risk $p_0$ rises with X. The odds ratio for G moves in the opposite direction for the same reason, falling by roughly a third between X = −2 and X = +2 SD (Figure 3B). Neither summary is invariant to baseline risk, so a variant with fixed causal structure and a variant whose effect genuinely varies with context can produce the same reported penetrance and the same stratum-specific odds ratio.

PN does not resolve this at any single stratum. What separates the two cases is the profile of PN across X. PN declines with X because the share of affected carriers for whom the variant was necessary falls as competing causes accumulate.

Recovering that profile requires enough affected carriers in every stratum for the stratum-specific estimates to separate. PN is an individual-level estimand estimated by a stratum-level average, so its precision is governed by case counts within strata rather than by total sample size, and the tails of X are the constraint. This is a second structural reason, alongside the ascertainment argument, to calibrate PN in population-based cohorts rather than clinic-based series.

**PMN**

The mediator M is itself a binary outcome of G, and an analogous monotonicity assumption M(1) ≥ M(0) defines three latent mediator response types ($U_M$) by their joint potential mediator values {M(0), M(1)} (Table S7).

**Table S7. Mediator response types ($U_M$) , {M(0)=1, M(1)=0} is excluded by monotonicity.**

| Type | M(0) | M(1) | Interpretation |
|---|---|---|---|
| M-Never | 0 | 0 | Biomarker absent regardless of G |
| M-Caused | 0 | 1 | Biomarker present only when G present |
| M-Always | 1 | 1 | Biomarker constitutively elevated |

The outcome response types ($U_Y$) are then defined conditional on the mediator pathway, yielding four types that capture whether disease depends on the biomarker, on G directly, on both, or on neither. The full latent type space is the cross-classification of $U_M$ and $U_Y$, producing 12 joint types (Table S8), one of which (M-Never × Y-M-only) is effectively 0 by construction because biomarker-dependent disease cannot occur in individuals whose biomarker is constitutively absent. The truth PMN is then the population fraction of jointly Necessary types i.e. those for whom Y(0,M(0))=0, among biomarker-positive affected carriers (G=1, M=1, Y=1).

**Table S8. Latent causal types defined by cross-classification of mediator response type (U_M) and outcome response type (U_Y)**

| $U_M$ | $U_Y$ | M(0) | M(1) | Y(0) | Y(1) | Contributes to PN? | Contributes to PMN? | Interpretation |
|---|---|---|---|---|---|---|---|---|
| M-Never | Y-Never | 0 | 0 | 0 | 0 | No | No | No disease, no biomarker |
| M-Never | Y-M-only | — | — | — | — | — | — | M never elevated, M-only disease impossible |
| M-Never | Y-G-only | 0 | 0 | 0 | 1 | Yes | No | Direct G effect; biomarker uninvolved |
| M-Never | Y-Always | 0 | 0 | 1 | 1 | No | No | Phenocopy; no biomarker |
| M-Caused | Y-Never | 0 | 1 | 0 | 0 | No | No | Biomarker caused by G; disease absent |
| M-Caused | Y-M-only | 0 | 1 | 0 | 1 | Yes | Yes | Full mediated path G→M→Y |
| M-Caused | Y-G-only | 0 | 1 | 0 | 1 | Yes | Yes | Direct G effect; biomarker also caused by G |
| M-Caused | Y-Always | 0 | 1 | 1 | 1 | No | No | Phenocopy; biomarker caused by G |
| M-Always | Y-Never | 1 | 1 | 0 | 0 | No | No | Constitutive biomarker; no disease |
| M-Always | Y-M-only | 1 | 1 | 1 | 1 | No | No | Disease from constitutive biomarker regardless of G |
| M-Always | Y-G-only | 1 | 1 | 0 | 1 | Yes | Yes | Direct G effect; constitutive biomarker |
| M-Always | Y-Always | 1 | 1 | 1 | 1 | No | No | Phenocopy; constitutive biomarker |

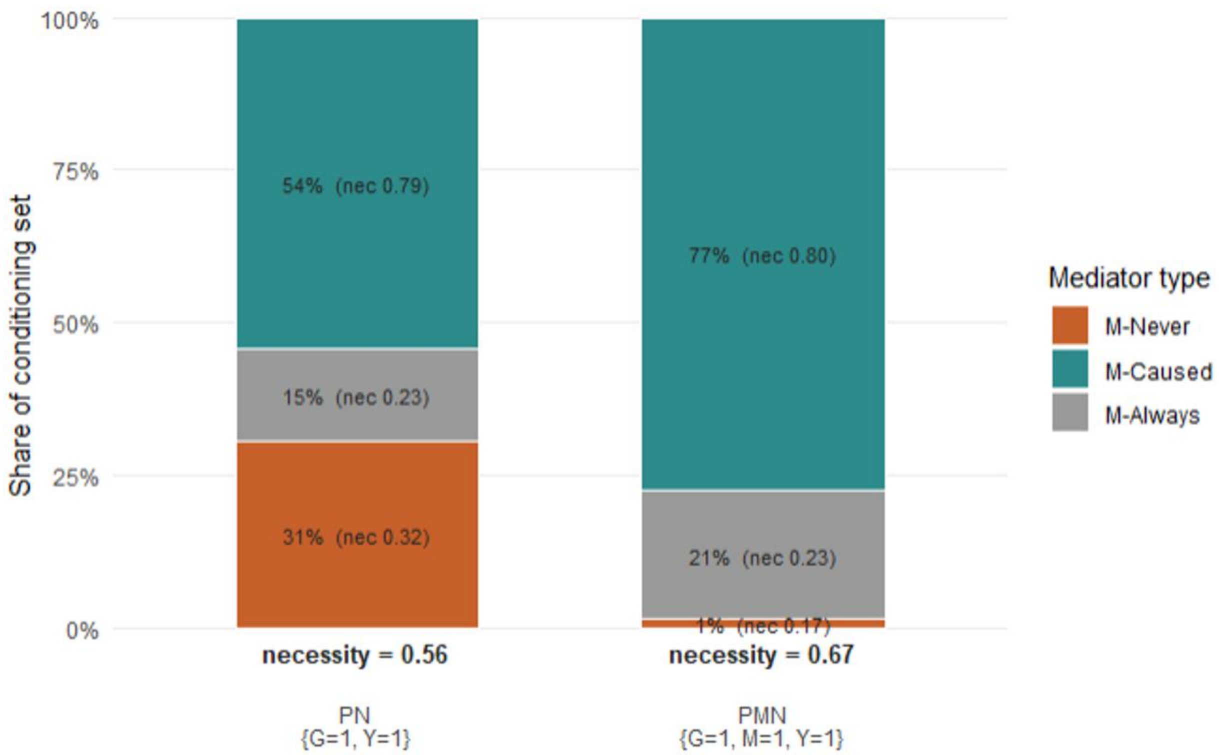


**Figure S2. Causal type composition in the {G,M,Y} model.** Stacked bars give the share of each conditioning set contributed by the three mediator response types defined in Table S7: M-Never (orange), M-Caused (teal) and M-Always (grey). The left bar is the PN conditioning set, affected carriers {G = 1, Y = 1}; the right bar is the PMN conditioning set, affected biomarker-positive carriers {G = 1, M = 1, Y = 1}. Within each segment the first value is the share of the conditioning set and the value in parentheses is the mean individual necessity of that type, defined as P(Y(0) = 0) for the PN set and P(Y(0, M(0)) = 0) for the PMN set, computed from known type membership. The bold value beneath each bar is the share-weighted mean of the segment necessities, which equals oracle PN and oracle PMN respectively. Single simulated population at baseline parameters (M-Never Always-type mass s = 10%, background disease rate 5%, n = [N], seed [S]); no Monte Carlo replication.

#### Effect of Uncertainty in the Data Generating Process (DGP) - Two-test sequential protocol

PMN is arithmetically defined for any dataset with observable ($G$, $M$, $Y$) triplets, but the formula does not internally verify that $M$ operates as a mediator of the $G \rightarrow Y$ relationship rather than as a proxy or independent cause. Two empirical preconditions are required before PMN can be interpreted as mediated causal necessity. The a-path test evaluates OR($G$, $M$) in an unselected population cohort: a non-null association confirms that the variant causes the biomarker, establishing the $G \rightarrow M$ link. The b-path test evaluates OR($M$, $Y \mid G = 0$) among non-carriers only: restricting to $G = 0$ removes the variant's direct effect on $Y$, so the association reflects only $M$'s independent causal contribution to $Y$, establishing the M → Y link. Both tests must pass for PMN to be interpretable as mediated necessity. The three-DGP matched simulation (Figure S3) demonstrates this discriminating capacity: three data-generating processes that share identical P($Y = 1 \mid G$, $M$) cell probabilities, and therefore identical observational distributions, differ in latent causal structure, and the two-test protocol correctly classifies each. Both the a-path OR(G,M) and b-path OR(M,Y|G=0) map directly to observational data: the a-path is estimated from the G–biomarker association in population biobanks; the b-path is estimated from epidemiological studies of the biomarker as a risk factor or predictor.

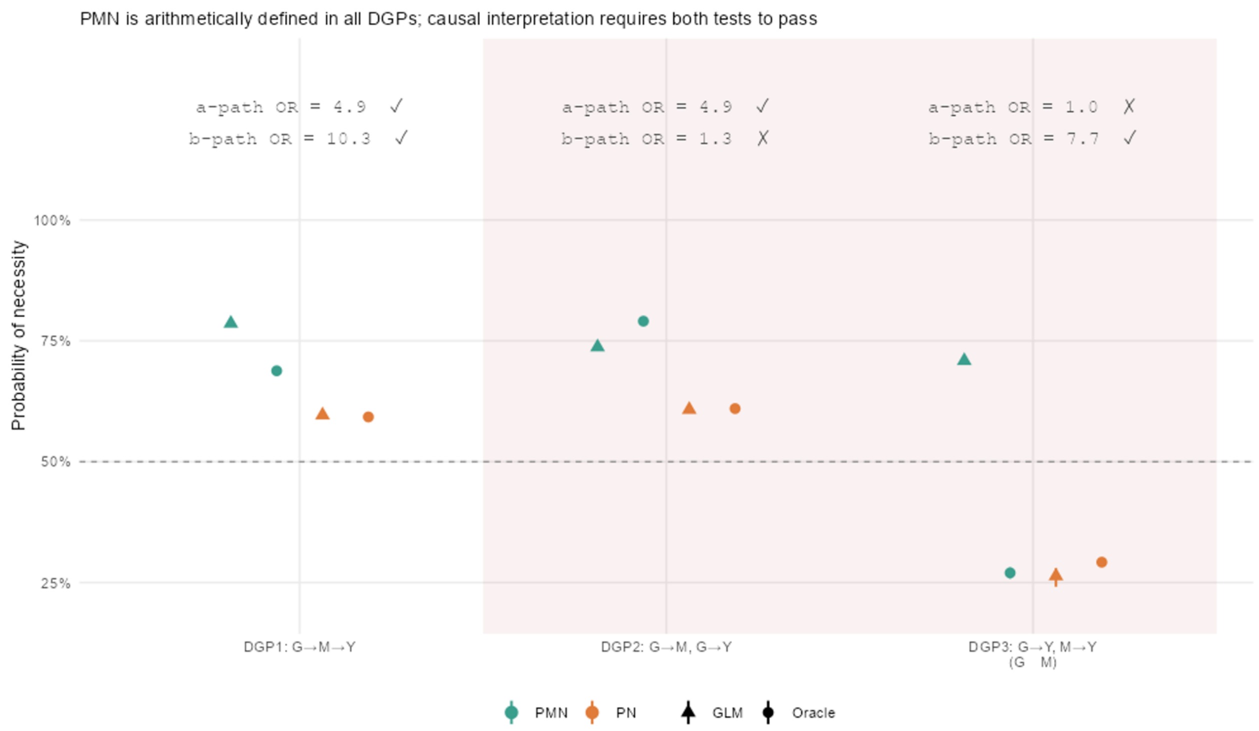


**Figure S3. Comparison of Three Possible Data Generating Processes.** Three data generating processes relating to G,M,Y are shown. We use two tests to probe the causal relationships: a-path test - OR(G, M) in the unselected cohort. Passes at OR > 2; b-path test - OR(M, Y | G = 0), among non-carriers only. (restricting to G = 0 removes the variant's direct effect on Y, so any residual M–Y association reflects M's own contribution); passes at OR > 2. In DGP1 the variant causes the biomarker (both a-path and b-path test pass); PMN is identified and carries the intended interpretation of mediated causal necessity. In DGP2 M is a proxy for G but has no effect on Y (a-path test passes, b-path fails). In DGP 3 G and M are independent causes of Y (a-path fails, b-path passes). Although DGP2 and DGP3 PMN is arithmetically defined it has no causal interpretation (pink shaded area).

### Cross World Independence Assumption

Consider the canonical mediator graph G → M → Y: the cross-world independence assumption posits that the outcome Y(0,m) (under treatment G=0 and a fixed mediator level M=m) is independent of the counterfactual mediator value M(0) (under treatment level G=0). While standard within-world independence (no unobserved confounding) is often sufficient for total effects, mediation requires bridging two hypothetical scenarios: the 'intervention world' where we set G to 1 and the alternate world where G is 0 (and the latter determines the level of the mediator without G intervention). By assuming $Y(0,m) \perp M(0)$, we rule out the existence of reciprocal confounding or "cross-world" correlation. Specifically, it implies that there are no variables that act as a consequence of the treatment G=1 which then go on to confound the relationship between the mediator M and the outcome Y. Without this assumption, the "natural" level of the mediator M(0) could carry information about individual-level latent characteristics that would also affect Y(0, m), making it impossible to disentangle the indirect path from the direct path using purely observational or even standard experimental data.

In our PO model of single gene effects the statement $Y(0,m) \perp M(0)$ is credible. Given G is a germline variant, it is fixed at conception. The counterfactual M(0) distribution is plausibly independent of the potential outcome Y(0,m) conditional on confounders. The cross-world independence assumption (A7: $Y(0,m) \perp M(0) \mid Z$) is the most demanding identification condition in the PMN analysis and is not reducible to standard no-unmeasured-confounding language.

**Crossworld Assumption in Inschemic Heart Disease Example**

Assumption A7 requires that the counterfactual outcome Y(0,m) (the IHD status a person would have if both the *LDLR* variant and its effect on the mediator were absent) is independent, given Z, of M(0) (the hypercholesterolemia status the same person would have in the absence of the variant). These potential outcomes come from two distinct hypothetical worlds, making A7 in principle untestable. In analyses of modifiable exposures, A7 is characteristically threatened by post-baseline confounders: variables that arise after exposure assignment, affect the mediator, and independently influence the outcome. Germline variant status substantially attenuates this concern: G precedes all subsequent biological processes, so time-varying confounding, behavioral feedback, and selective exposure, the mechanisms that most commonly generate A7 violations, have no analogue in this setting.

The residual concern specific to this application is an unmeasured variable U that drives hypercholesterolemia through non-*LDLR* pathways, for example, rare variants in PCSK9 or APOB, dietary lipid intake, insulin resistance, which independently elevate IHD risk. Such a U would generate positive correlation between M(0) and Y(0,m=1) among non-carriers. Individuals whose hypercholesterolemia is attributable to non-*LDLR* mechanisms also face elevated IHD risk through those same pathways. Under positive correlation, the identification formula underestimates E[Y(0,M(0))] by omitting the covariance term, and PMN is consequently overstated. The CAD polygenic risk score included as a covariate in Z captures a substantial share of the polygenic contribution to both M and Y, attenuating this concern, but rare lipid-pathway variants and non-genetic confounders are not fully absorbed. Formal sensitivity bounds for A7, analogous to the E-value approach for unmeasured confounding or the sensitivity parameters developed for the no-$\{M \to Y\}$-confounding assumption in the broader mediation literature, have not been computed here. The bin-level PMN analysis is designed precisely for this purpose: it provides stratum-level PN and PMN estimates with bootstrap confidence intervals that a clinician might use to characterize causal certainty in broad terms, without requiring the individual-level scoring precision that formal A7 sensitivity bounds would enable.

### Variant Binning for Rare Variant Analysis at an Example Locus

#### Table S9 AF Tier (F axis)

| Tier | Label | gnomAD NFE AF Range | Genetic Principle | Practical Notes |
|---|---|---|---|---|
| F1 | Ultra-rare | < 0.01% (< 0.0001) | Strong purifying selection signal; consistent with high penetrance Mendelian variant | Variants absent from gnomAD assigned F1 with gnomad_absent flag |
| F2 | Rare | 0.01–0.1% (0.0001–0.001) | Rare enough for moderate causal prior; some population-specific founder variants | AF should be ancestry-matched; NFE default, override per individual |
| F3 | Low-frequency | 0.1–1% (0.001–0.01) | Weaker causal prior; more likely represents low-penetrance or risk allele | PN estimates expected to be lower; wider CIs due to carrier count dilution |
| EXCLUDE | Common | ≥ 1% (≥ 0.01) | BA1 equivalent; frequency incompatible with high-penetrance dominant disease causation | Retained in variant file with flag; not entered into PN estimation |
| ABSENT | Novel | AF = 0 or not in gnomAD | Ultra-rare by definition; assigned F1 | Strongest case for bin-level PN inheritance — no prior observation available |

#### Table S10 Consequence Tier (T axis)

| Tier | Label | Variant Classes | Score Criteria | Genetic Principle | Practical Notes |
|---|---|---|---|---|---|
| T1 | High-confidence LoF | stop_gained, frameshift, canonical splice ±1/2, start_lost, stop_lost | None — consequence sufficient | LoF in a haploinsufficient gene is the strongest prior for causal necessity; PVS1-equivalent | Requires ClinGen gene-level LoF mechanism flag; LDLR qualifies cleanly |
| T2 | Strong missense, concordant | missense, inframe indel | ≥2 votes from: REVEL ≥ 0.75, AM ≥ 0.80, ESM1b ≤ −7.5 | Concordance across independent computational metrics reduces false positive rate | Discordant single-vote variants separated into T2_disc for sensitivity analysis |
| T2_disc | Strong missense, discordant | missense, inframe indel | Exactly 1 strong vote | One metric predicts strong deleteriousness; others do not support it | Use as T3 in primary analysis; sensitivity stratum |
| T3 | Moderate missense | missense, inframe indel | ≥1 moderate vote: REVEL 0.50–0.75, AM 0.56–0.80, ESM1b −7.5 to −5.0; CADD ≥ 20 as fallback | Computational support present but below concordance threshold | CADD used only when REVEL/AM/ESM1b absent — documented as fallback |
| T4 | Weak / synonymous | synonymous, other | No criteria met | Minimal prior for causal capacity; functions as internal negative control in PN estimation | PN should be near zero; deviation signals bin contamination or population stratification artifact |
| T_splice | Predicted splice-disrupting | Any SO term | SpliceAI max delta ≥ 0.50 | SpliceAI captures cryptic and deep intronic splice variants missed by consequence annotation | Override tier — applied regardless of SO term; treat as T1-equivalent when splicing is gene mechanism |

#### Table S11 Score Voting Logic

| Score | Strong Vote Threshold | Moderate Vote Threshold | Direction | Rationale |
|---|---|---|---|---|
| REVEL | ≥ 0.75 | 0.50–0.75 | Higher = more deleterious | Ensemble score; well validated against clinical variant sets |
| AlphaMissense | ≥ 0.80 | 0.56–0.80 | Higher = more deleterious | Protein LM score; used as continuous deleteriousness metric — class labels not used |
| ESM1b | ≤ −7.5 | −7.5 to −5.0 | More negative = more deleterious | Evolutionary protein LM; captures signal when alignment depth limits REVEL |
| CADD phred | N/A | ≥ 20 | Higher = more deleterious | Fallback only when REVEL/AM/ESM1b all absent; never votes for T2 |
| SpliceAI max delta | ≥ 0.50 (override) | — | Higher = more disruptive | Overrides consequence tier independently; not part of vote count |

**T2 requires ≥ 2 strong votes. T2_disc = exactly 1. T3 = ≥ 1 moderate vote with 0 strong votes.**

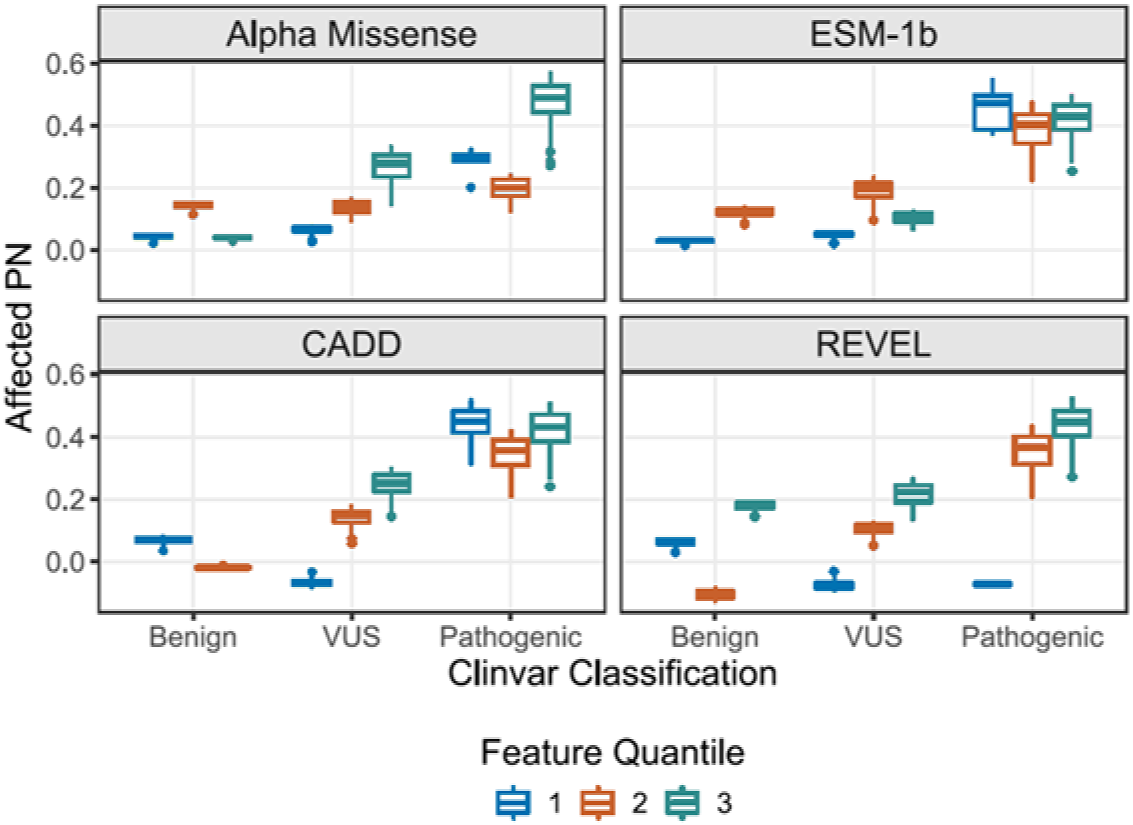


**Figure S4 Consistency of Computational Deleteriousness Scores.** Box plots show Affected PN stratified by ClinVar classification and feature quantile (Q1–Q3) for each score, demonstrating monotonic concordance between computational deleteriousness rankings and estimated probability of necessity across classification categories.
